# Modelling interventions to control COVID-19 outbreaks in a refugee camp

**DOI:** 10.1101/2020.07.07.20140996

**Authors:** R. Tucker Gilman, Siyana Mahroof-Shaffi, Christian Harkensee, Andrew T. Chamberlain

**Author notes:** **Correspondence to:** R. Tucker Gilman, Centre for Crisis Studies and Mitigation, University of Manchester, Manchester, M13 9PL, UK.

## Abstract

Refugee camp populations are expected to be vulnerable to COVID-19 due to overcrowding, unsanitary conditions, and inadequate medical facilities. Because there has been no COVID-19 outbreak in a refugee camp to date, the potential for nonpharmaceutical interventions to slow the spread of COVID-19 in refugee camps remains untested. We used an agent-based model to simulate COVID-19 outbreaks in the Moria refugee camp, and we studied the effects of feasible interventions. Subdividing the camp (’sectoring’) “flattened the curve,” reducing peak infection by up to 70% and delaying peak infection by up to several months. The use of face masks coupled with efficient isolation of infected individuals reduced the overall incidence of infection and sometimes averted epidemics altogether. These interventions must be implemented quickly to be effective. Lockdowns had little effect on COVID-19 dynamics. Our findings provide an evidence base for camp managers planning intervention strategies against COVID-19 or future epidemics.

## Introduction

There are more than 70 million refugees and internally displaced persons worldwide, including more than 20 million living in displacement camps.^1^ Displaced populations are expected to be vulnerable to COVID-19 due to poor nutrition, high rates of pre-existing disease, and inadequate access to healthcare.^2-5^ COVID-19 may spread rapidly in camps due to overcrowding, poor sanitation, and frequent close contact among residents (*e.g*., in food lines, at shared toilets, and at shared washing facilities).^5-7^ Truelove and colleagues used a computational simulation to study a potential COVID-19 outbreak in a population modelled on the Kutupalong-Balukhali refugee camp in Bangladesh, and estimated that up to 98% of the population could become infected over a short period, overwhelming the camp’s limited medical facilities.^8^ Many countries have imposed interventions such as mandatory social distancing, isolation of confirmed cases, or general lockdowns to slow the spread of COVID-19, and in some cases these have been successful.^9-11^ However, whether similar interventions can be effective in the uniquely challenging setting of a displacement camp is unknown.^7^

The Moria refugee camp on the island of Lesbos, Greece, is Europe’s largest displacement camp. A former prison, it was converted into a refugee reception facility with the arrival of people fleeing the Syrian civil war in 2015. Designed to hold 3,000 people, it now houses nearly 20,000 in an area of less than 1 km^2^.^12^ Non-governmental organizations (NGOs) working in Moria report severe overcrowding, poor sanitation, a lack of hygiene facilities (*e.g*., toilets, showers, 24-hour running water), and queuing at central facilities (*e.g*., food lines).^13,14^ The population has little access to healthcare outside the camp, and there is a lack of adequate healthcare in the camp (*e.g*., no 24-hour service, provided only by volunteer organizations). Approximately 5% of the camp’s population is highly vulnerable to COVID-19 infection, including people with chronic health conditions and those over 65 years of age. COVID-19 has not yet reached the camp. However, there have been cases of COVID-19 on Lesbos,^13,15^ and the virus is likely to reach the camp soon.

Because there has not yet been a COVID-19 outbreak in a displacement camp, there is no empirical data to show which interventions are best able to combat the spread of the disease in this setting. In the absence of empirical data, mathematical and computational models can provide an evidence base for managers planning intervention strategies, and lessons learned may be transferable to other displacement camps.

Displacement camp populations are spatially structured. In Moria, residents interact most frequently with other members of their own households. They interact with members of nearby households during daily activities or at shared toilet facilities, and they interact with residents from all parts of the camp at the camp’s single shared food line. This interaction structure will affect how COVID-19 spreads through the camp, and interventions that change the interaction structure may alter the trajectory of outbreaks. Previous modelling of COVID-19 outbreaks in displacement camps used compartmental models,^8^ which assume that populations are well-mixed. Agent-based models that track individuals through simulated daily movements are better able to capture transmission dynamics in structured populations.^16^ Furthermore, agent-based models can easily simulate interventions that change interaction structures in populations, which is more difficult to achieve with compartmental models.

We developed the first agent-based model of an epidemic in a displacement camp setting and applied it to evaluate potential interventions to combat the spread of COVID-19 (see Methods). The model tracks individuals as they undertake daily activities in a simulated camp. COVID-19 can be transmitted when infected and susceptible individuals interact. The demographics and spatial structure of the model camp simulate Moria, and the parameters that control COVID-19 transmission rates and disease progression are estimated from the literature.^17-28^ We modelled COVID-19 outbreaks without interventions and in the presence of four interventions feasible for displacement camps: i) sectoring, ii) transmission reduction, iii) remove-and-isolate, and iv) lockdown. In sectoring, the central food line is eliminated and the camp is divided into sectors. Each sector has its own food line, and each individual uses the food line in the sector in which it lives. Thus, time spent in the food line is reduced and transmission in the food line becomes local rather than global. Transmission reduction could be any policy or behaviour that reduces the probability of transmission when individuals interact (*e.g*., the use of face masks, frequent hand washing, maintaining safe distances from others). Frequent hand-washing and maintaining safe distances from others are likely to be impossible in Moria^5^, but residents have been provided with face masks. Therefore, we simulated face mask use, and we refer to this intervention as “face masks” hereafter. In remove-and-isolate, households with symptomatic individuals are moved to an isolation facility to prevent onward transmission of the infection. Finally, in lockdown, individuals are constrained to remain within some distance of their homes, except when visiting shared toilets or food lines. A small proportion of the population violates the lockdown rule. For each intervention or combination of interventions, we conducted simulations in which we introduced an infected individual into the population, and we recorded the proportion of times that an epidemic occurred (*i.e*., 20 or more people became infected). If an epidemic occurred, we recorded i) the peak proportion of the population infected, ii) the time from the introduction of the first case until peak infection, and iii) the total proportion of the population that became infected. For remove-and-isolate interventions, we also recorded the maximum number of individuals kept in isolation to assess the feasibility of the intervention.

## Results

The probability that COVID-19 is transmitted when individuals interact is poorly understood. Therefore, we studied both low- and high-transmission scenarios based on low-^19,22^ and high-end^21,25^ estimates from the literature (see Methods). The true transmission probabilities for COVID-19 are likely to fall between these estimates. How individuals use space and interact with others in Moria or other refugee camps has not been studied. Therefore, we modelled low- and high-movement scenarios and low- and high-interaction scenarios based on estimates provided by healthcare workers in Moria. In the body of this paper we present results for the low-movement, high-interaction scenario, but results are qualitatively similar for other scenario combinations (supplementary tables S1-S11). To simulate face mask use, we reduced the odds of transmission by a factor of 0.32 in all interactions outside the home. A similar reduction has been achieved with the use of surgical masks for other respiratory viruses.^23^ The other interventions can be imposed with different intensities (*e.g*., more or fewer sectors, more or less efficient detection of symptoms in remove-and-isolate interventions, stricter or less strict lockdowns). In the body of the paper we present results for sectoring into 16 sectors, remove-and-isolate with a daily detection rate of 50% for symptomatic individuals, and lockdowns where individuals are required to stay within 10 m of their homes and 10% of the population violates the lockdown rule. We present qualitatively similar results for interventions with other intensities in supplementary tables S3-S8.

In the absence of interventions, the introduction of a single COVID-19 case into the model population almost always (≥97%) led to epidemics in both the low- and high-transmission scenarios (tables 1, S1). In the low-transmission scenario, the median peak infection included 67%) of the population and occurred 55 days after the index case appeared (figure 1 A). In the high-transmission scenario, the median peak infection included 98% of the population and occurred on day 25 (figure 1B). In total, 98% and >99% of the population became infected in the low- and high-transmission scenarios, respectively (table 1).

**Figure 1.**
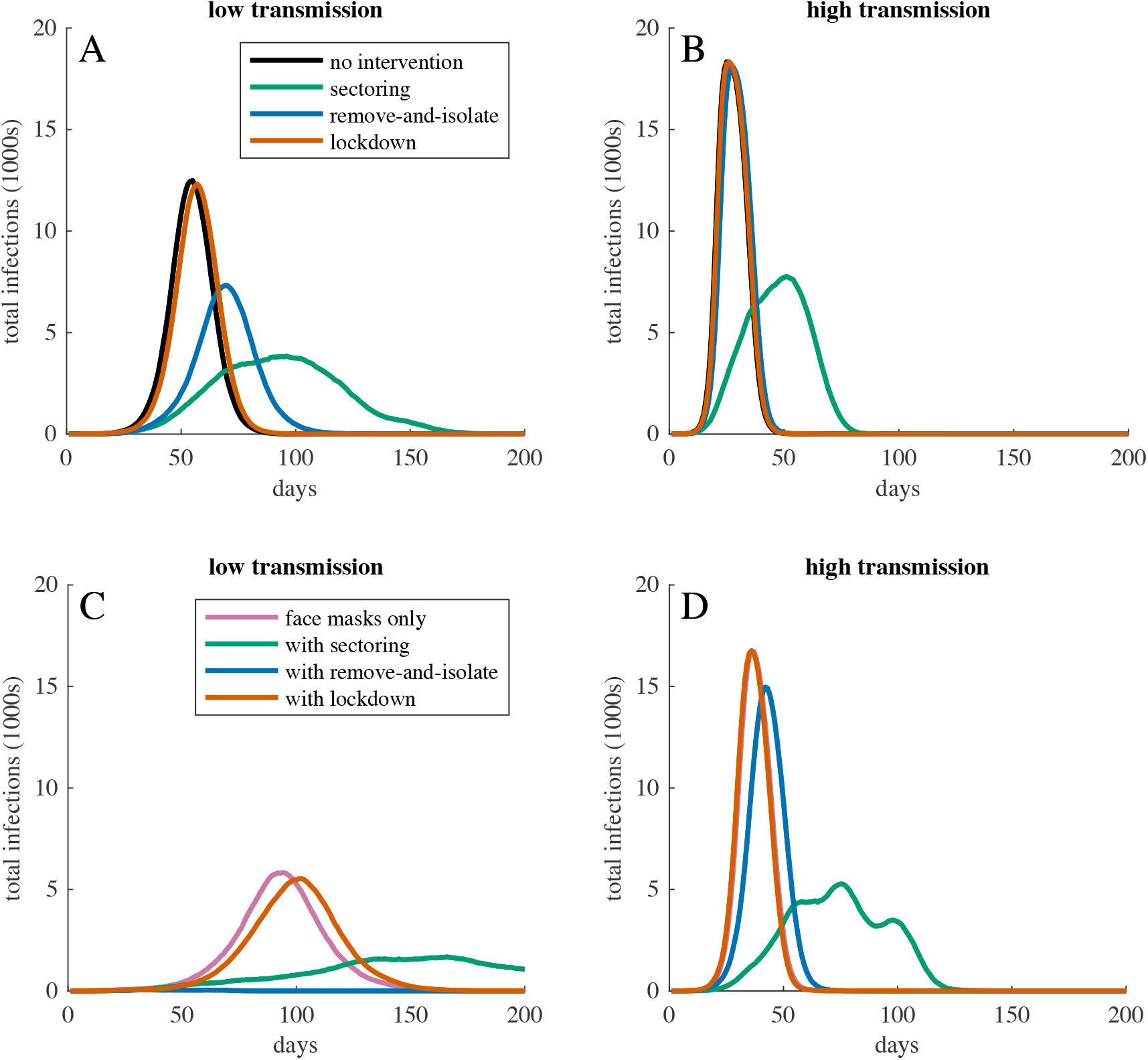
Total infections over time for COVID-19 epidemics with different interventions in populations with low movement, high interaction, and (A,C) low or (B,D) high transmission probabilities. Panels (A,B) show dynamics without face mask use, and (C,D) show dynamics with face mask use. Curves show the most representative simulation (*i.e*., the simulation with the peak infection and peak infection date closest to the median) for the corresponding intervention. When transmission probabilities were high (B,D), only sectoring meaningfully reduced or delayed peak infection. When transmission probabilities were sufficiently low (*i.e.*. low transmission with face masks, C), remove-and-isolate interventions were able to stop epidemics. In (D), the line for face mask use only is concealed behind the line for face masks with lockdown.

**Table 1.**
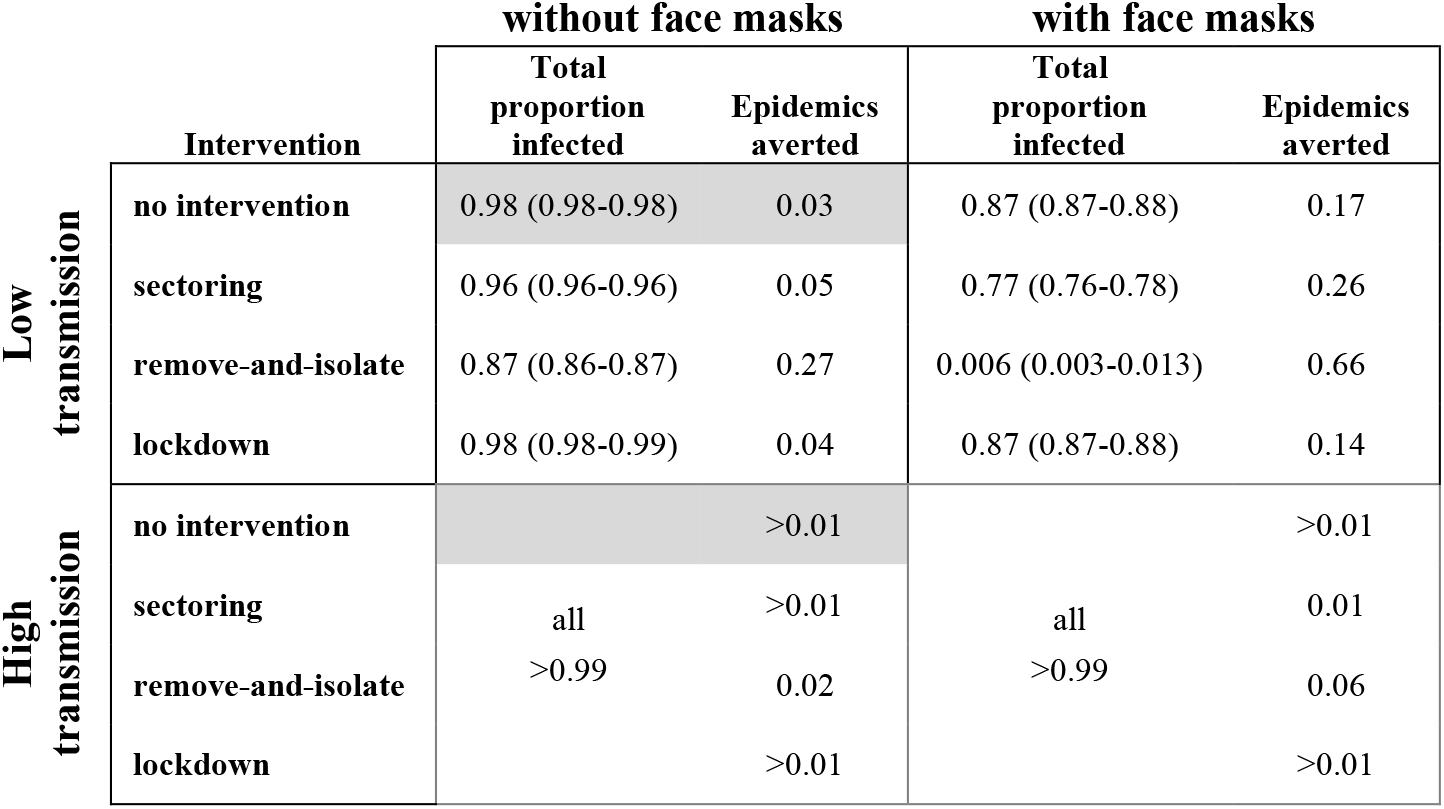
Total proportion of the population infected and epidemics averted without or with interventions in the low- and high-transmission scenarios. For total proportions infected, we report medians and interquartile ranges for all simulations in which epidemics occurred. For epidemics averted, we report proportions of 200 simulations. Grey cells indicate simulations without interventions.

Interventions were able to slow or stop the spread of COVID-19 (figure 1; tables 1, S2-S11). Sectoring reduced and delayed the peak infection in both the low-transmission (median peak infection 20% on day 98) and high-transmission (median peak infection 41% on day 51) scenarios, but most individuals ultimately became infected (low-transmission scenario: total infection 96%, epidemics averted 5%; high-transmission scenario: total infection >99%, epidemics averted <1%). Face masks reduced and delayed the peak infection in the low-transmission scenario (median peak infection 31% on day 96) but were less effective in the high-transmission scenario (median peak infection 90% on day 36) (figure 1C,D). In the low-transmission scenario, face masks also reduced the proportion of the population that became infected (total infection 87%, epidemics averted 17%). In the low-transmission scenario, remove-and-isolate interventions averted 27% of epidemics. When epidemics occurred, remove-and-isolate interventions reduced and delayed the peak infection, but required concurrently isolating more than half of the population (table S4). In the high-transmission scenario, remove-and-isolate interventions had little effect on epidemics (median peak infection 97% on day 27). Lockdowns had little effect on epidemics in either the low-transmission (median peak infection 66% on day 57) or high-transmission (median peak infection >98%) on day 26) scenarios.

The use of face masks augmented the effects of sectoring and remove-and-isolate interventions (figure 1C,D; tables 1, S6-S8). In the low-transmission scenario, sectoring combined with face masks reduced the median peak infection to 9% on day 167, limited total infection to 77% of the population, and averted 26% of epidemics. In the high-transmission scenario, sectoring combined with face masks reduced the median peak infection to 28% of the population on day 76, but >99% of the population eventually became infected. In the low-transmission scenario, remove-and-isolate combined with face masks prevented most epidemics (median peak infection 0.2%, total infection 0.6%, 66% of epidemics averted). However, in the high-transmission scenario, remove-and-isolate combined with face masks was little better than face masks alone. Similarly, in both scenarios, lockdown combined with face masks was little better than face masks alone.

Sectoring and remove-and-isolate interventions helped control epidemics, but had to be implemented early to be maximally effective (figure 2; tables S9, S10). If face masks were used but sectoring was not implemented until 1% of the population showed symptoms in the low-transmission scenario, then the median peak infection increased from 9% to 19% and the proportion of epidemics averted dropped from 26% to 14%. In the high-transmission scenario, peak infection increased from 28% on day 76 to 78% on day 38. If remove-and-isolate was not implemented until 1% of the population showed symptoms in the low-transmission scenario, then the median peak infection increased from 0.2% to 8.6%, the median total infection increased from 0.6% to 30%, and epidemics averted dropped from 66% to 10%. In the high-transmission scenario, remove-and-isolate was not effective even if it was implemented early (figure 1D).

**Figure 2.**
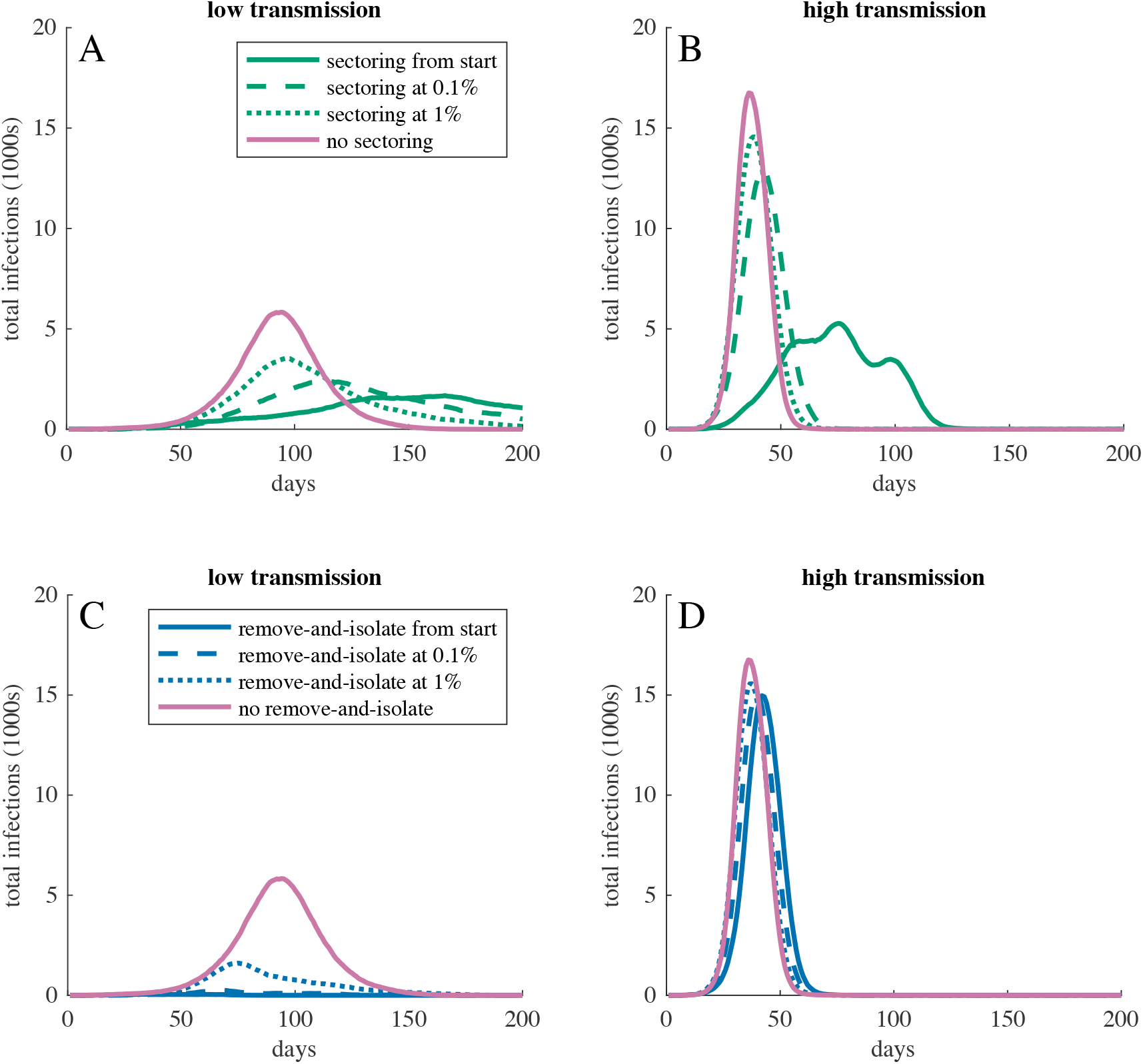
Total infections over time for COVID-19 epidemics when (A,B) sectoring or (C,D) remove-and-isolate interventions started before the virus arrived, when 0.1% of the population had symptoms, when 1% of the population had symptoms, or not at all. Face masks were in use throughout all simulations. (A,C) show the low-transmission and (B,D) show the high-transmission scenario. Curves show the most representative simulation for the corresponding intervention. In all cases, a delayed start to the intervention resulted in higher peak infection. In the high-transmission scenario, even a slightly delayed start eliminated most gains that could be achieved by the intervention.

## Discussion

Displacement camp populations are expected to be vulnerable to COVID-19 and other epidemics due to poor sanitation, crowded conditions, high rates of pre-existing disease, and inadequate access to healthcare.^2,3,6^ Without intervention, a single case of COVID-19 introduced into our model population almost always led to a severe epidemic that rapidly spread through the entire population. Sectoring, remove-and-isolate interventions, and the use of face masks slowed the spread of infection, and in some cases stopped epidemics altogether. These interventions must be implemented early to be maximally effective. Our results can help displacement camp managers choose the most effective interventions to protect vulnerable populations from COVID-19 and other epidemics.

Dividing the camp into sectors with separate food lines reduced and delayed the infection peak. Reducing the number of people that are infected at the same time may help alleviate pressure on limited medical services both in the camp and in the surrounding community.^10^ Our model assumes that sectoring can prevent meetings, and thus transmission, between individuals from different parts of the camp. If this is not true, then sectoring may be less effective than our model suggests. Furthermore, while sectoring slowed the rate at which epidemics spread through the camp, it rarely averted epidemics altogether and had only a small effect on the total number of individuals that became infected. Thus, while sectoring may reduce pressure on medical services, sectoring alone is unlikely to protect vulnerable members of the population who may be at heightened risk due to COVID-19 with or without medical attention.

In contrast to sectoring, both face mask use and remove-and-isolate interventions reduced the total number of people that became infected. When infectiousness was at the low end of published estimates for COVID-19, face masks coupled with an efficient remove-and-isolate plan prevented more than 65% of COVID-19 introductions from becoming epidemics and limited the median total infection to <1% of the population. Combining these interventions with sectoring produced further gains. However, the effectiveness of face masks and remove-and-isolate interventions was sensitive to the infectiousness of the virus. If the infectiousness was at the high end of published estimates, face mask use and remove-and-isolate interventions had little effect on epidemics. Furthermore, remove-and-isolate interventions require that managers are able to quickly and accurately detect COVID-19 cases, and may be resource-intensive if they fail to avert epidemics completely. Because people must be maintained in isolation until managers are sure they are no longer infectious, the maximum number of people in isolation will usually be larger than the peak infection (supplementary tables S4, S7, S10, and S11). If managers’ capacity to remove and isolate infected individuals is overwhelmed, then remove-and-isolate interventions will fail.

In our model, requiring individuals to remain within a small radius of their homes had little effect on epidemics. Even during lockdowns, transmission continued at shared toilets and food lines. Moreover, the lockdowns we studied were ambitious. For results reported in the body of the paper, we assumed that 10% of individuals would violate the lockdown rule, but in the UK more than 25% of young women and more than 50% of young men admit to regularly violating lockdown rules^29^ and similar patterns have been reported in the US.^30^ Thus, it is not clear that lockdowns of the sort we modelled will be effective at combatting the spread of COVID-19 in refugee camps. However, the number of interactions that individuals engage in each day can affect the dynamics of epidemics (compare shaded to unshaded rows in supplementary tables S1-S11). Thus, encouraging camp residents to limit their daily interactions may be a viable tool for slowing epidemics.

Sectoring and remove-and-isolate interventions must be implemented from the beginning of an outbreak if they are to be maximally successful. If interventions are not in place when the virus arrives, the virus can rapidly spread to all parts of the camp. It then becomes very difficult to contain. Background rates of respiratory infection in displacement camps are high^3,4^, which may make new infections difficult to detect. Thus, population managers should be prepared to impose interventions at the first threat of epidemic.

Our results provide valuable guidance to displacement camp managers, who currently lack empirical evidence to support intervention planning. However, the parameter values that underlie our results are estimated with uncertainty. The transmission probabilities for COVID-19 are estimated from the literature, which is rapidly evolving. The parameter values that describe how individuals move and interact in the camp are estimated from consultation with camp medical staff, and empirical data to confirm these estimates do not exist for Moria or any other displacement camp. Different transmission probabilities, and to a lesser extent different interaction rates, within the plausible range of values result in very different epidemics. Until parameter values can be more accurately estimated, our model should not be used to make quantitative predictions about peak infection rates, times to peak infection, or proportions of epidemics averted. Some qualitative predictions of the model also depend on the parameter values. For example, in the low-transmission scenario, combinations of the interventions we modelled can stop the spread of COVID-19. In the high-transmission scenario, sectoring can slow the epidemic, but almost the entire population is eventually infected. Thus, in the high-transmission scenario, the removal and shielding of vulnerable individuals (*i.e*., those over 65 years old, or with pre-existing conditions^2,17,18^) may be the only intervention that saves lives. Per interaction transmission rates are notoriously difficult to estimate empirically, and interaction rates and networks among members of vulnerable populations have rarely been studied. These are key parameters in agent-based epidemiological models, and agent-based models are better than classical compartmental models at simulating the spread of disease in structured and heterogeneous populations.^16^ Thus, empirical work to estimate interaction rates and per interaction transmission probabilities may be of great value. Finally, our model assumes that individuals that have recovered from COVID-19 cannot be re-infected at least for the duration of the epidemic, and evidence to support this assumption is limited.^31^ As more empirical data on COVID-19 becomes available, our model can be updated to provide more accurate predictions.

The model we present here is the first attempt to evaluate potential interventions to control the spread of COVID-19 in a displacement camp. We focused on Moria because severe overcrowding, poor sanitation, and frequent centralised queuing make it a particularly challenging setting for disease control,^6-8^ and because the recent arrival of COVID-19 on Lesbos makes intervention planning for the camp an urgent priority.^13,15^ However, with modified parameter values, our model can be applied to evaluate potential interventions to combat COVID-19 or other transmissible diseases in other displacements camps or vulnerable populations (*e.g*., urban slums^7^).

Many uncertainties exist about how COVID-19 will affect refugee camp populations, and whether feasible interventions can mitigate these effects. It is not possible to evaluate interventions with well-controlled experiments, because it would be unethical to apply interventions in some populations and withhold them from others. In the absence of empirical data, agent-based simulations like those we present here may offer the best opportunity to assess potential interventions and to plan management strategies that could save human lives.

## Methods

### Overview

We developed a spatially explicit agent-based model to simulate COVID-19 epidemics unfolding in a model refugee camp over discrete timesteps that correspond to days. The infection starts in one individual, and is transmitted probabilistically among individuals as they interact during daily activities. We modelled epidemics with no interventions, and epidemics where interventions or combinations of interventions were used to reduce disease transmission. We compared the peak number of infected individuals, the time to peak infection, and the total number of individuals infected, with or without interventions.

The parameter values that describe the population and the camp simulate the Moria refugee camp on Lesbos, Greece. The parameter values that describe disease progression and transmission are drawn from the literature. The parameter values that describe individuals’ movements in the camp are heuristic, but our qualitative predictions hold under other reasonable sets of parameter values (supplementary tables S1-S11).

Throughout these methods, we used “Moria” to refer to the Moria refugee camp and “camp” to refer to the camp in our model. We used “person” or “people” to refer to the residents of Moria, and we used “individuals” to refer to individuals in the model population.

### The population

The model population comprises 18,700 individuals. Each individual is characterised by its age, sex, condition, and disease state. Condition describes whether an individual is healthy or has a pre-existing condition that increases the risk of severe infection or mortality from COVID-19 (*i.e*., hypertension, diabetes, cardiovascular disease, or chronic lung disease^2^). Each individual is assigned an age, sex and condition that matches a randomly selected person from the medical records of the Moria camp. These characteristics do not change over time. The disease state describes the progression of a COVID-19 infection in an individual, and therefore does change over time. The initial disease state for all individuals is “susceptible.”

### The camp

Each individual is a member of a household that occupies either an isobox or a tent. Isoboxes are prefabricated housing units with a mean occupancy of 10 individuals. Tents have a mean occupancy of 4 individuals. A total of 8,100 individuals occupies isoboxes and 10,600 individuals occupy tents. These correspond to the numbers of people occupying isoboxes and tents in Moria. The exact occupancy of each isobox or tent is drawn from a Poisson distribution, and individuals are assigned to isoboxes or tents randomly without regard to sex or age. This is appropriate because many people arrive at Moria travelling alone, and thus isoboxes or tents may not represent family units.

The camp covers a 1 × 1 (*e.g*., km) square (figure 3). Isoboxes are assigned to random locations in a central square that covers one half of the area of the camp. Tents are assigned to random locations in the camp outside of the central square. There are 144 toilets evenly distributed throughout the camp. Toilets are placed at the centres of the squares that form a 12 × 12 grid covering the camp. The camp has one food line. The position of the food line is not explicitly modelled.

**Figure 3.**
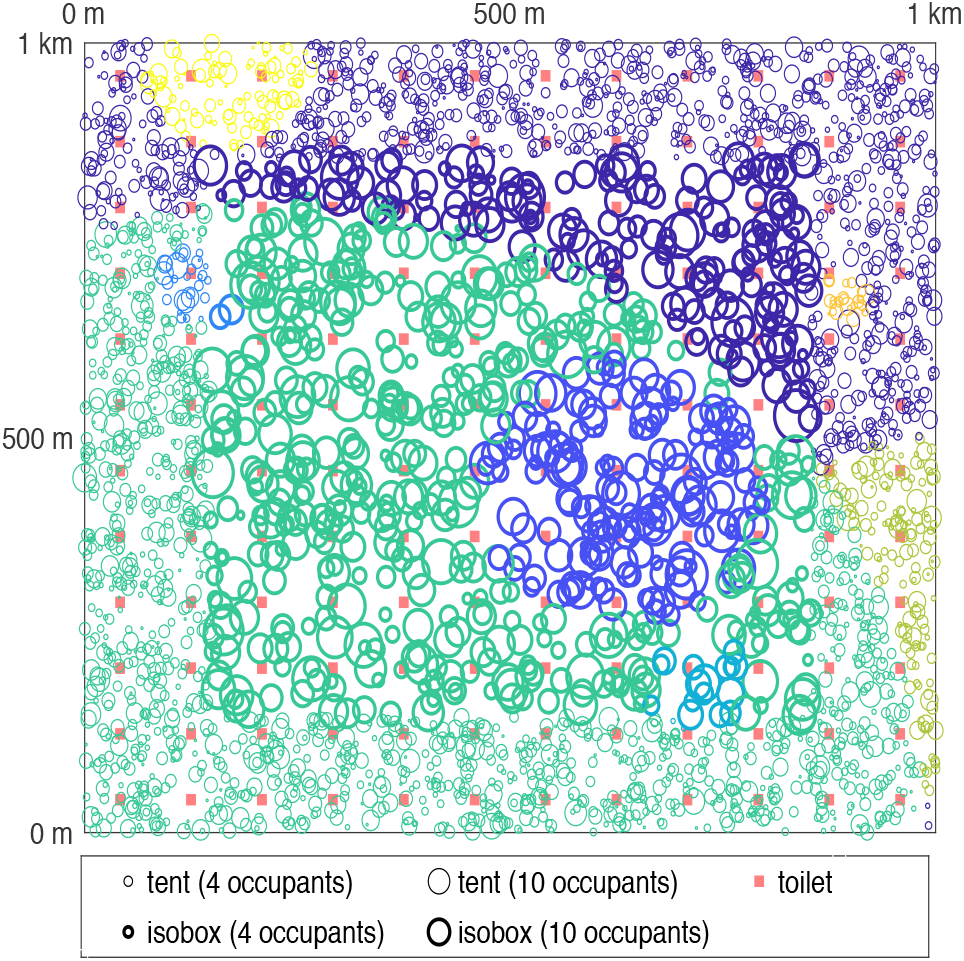
A representative example of the camp layout. Circles represent households. Circle size indicates household occupancy and circle colour indicates national or etliinc background. Isoboxes (bold circles) are centrally located, and tents (light circles) are peripherally located. Red squares represent toilets.

In Moria, the homes of people with the same ethnic or national background are spatially clustered, and people interact more frequently with others from the same background as themselves. To simulate ethnicities or nationalities in our camp, we assigned each household to one of eight “backgrounds” in proportion to the self-reported countries of origin of people in the Moria medical records. For each of the eight simulated backgrounds, we randomly selected one tent or isobox to be the seed for the cluster. We assigned the *x* nearest unassigned households to that background, where *x* is the number of households with that background. Thus, the first background occupies an area that is roughly circular, but other backgrounds may occupy less regular shapes (figure 3).

### Disease Progression

If an individual becomes infected, the infection progresses through a series of disease states (figure 4). The time from exposure until symptoms appear (*i.e*., the incubation period) is drawn from a Weibull distribution with a mean of 6.4 days and a standard deviation of 2.3 days.^28^ In the first half of this period, the individual is “exposed” but not infectious. In the second half, the individual is “pre-symptomatic” and infectious.^24^ Fractional days are rounded to the nearest whole day in discrete-time simulations. After the incubation period, the individual enters one of two states: “symptomatic” or “1^st^ asymptomatic.” Children under the age of 16 become asymptomatic with probability 0.836 and others become asymptomatic with probability 0.178.^20,26^ Individuals remain in the symptomatic or 1^st^ asymptomatic states for 5 days and are infectious during this period. After 5 days, individuals pass from the symptomatic to the “mild” or “severe” states with age- and condition-dependent probabilities following Verity and colleagues^17^ and Tuite and colleagues^18^. All individuals in the 1^st^ asymptomatic state pass to the “2^nd^ asymptomatic” state. Individuals are infectious in these states. On each day, individuals in the mild or 2^nd^ asymptomatic state pass to the recovered state with probability 0.37,^21^ and individuals in the severe state pass to the recovered state with probability 0.071.^27^ Recovered individuals are not infectious, and are not susceptible to reinfection. We did not model deaths explicitly, but this is unlikely to affect the dynamics of the epidemic if neither recovered nor dead individuals are infectious.

**Figure 4.**
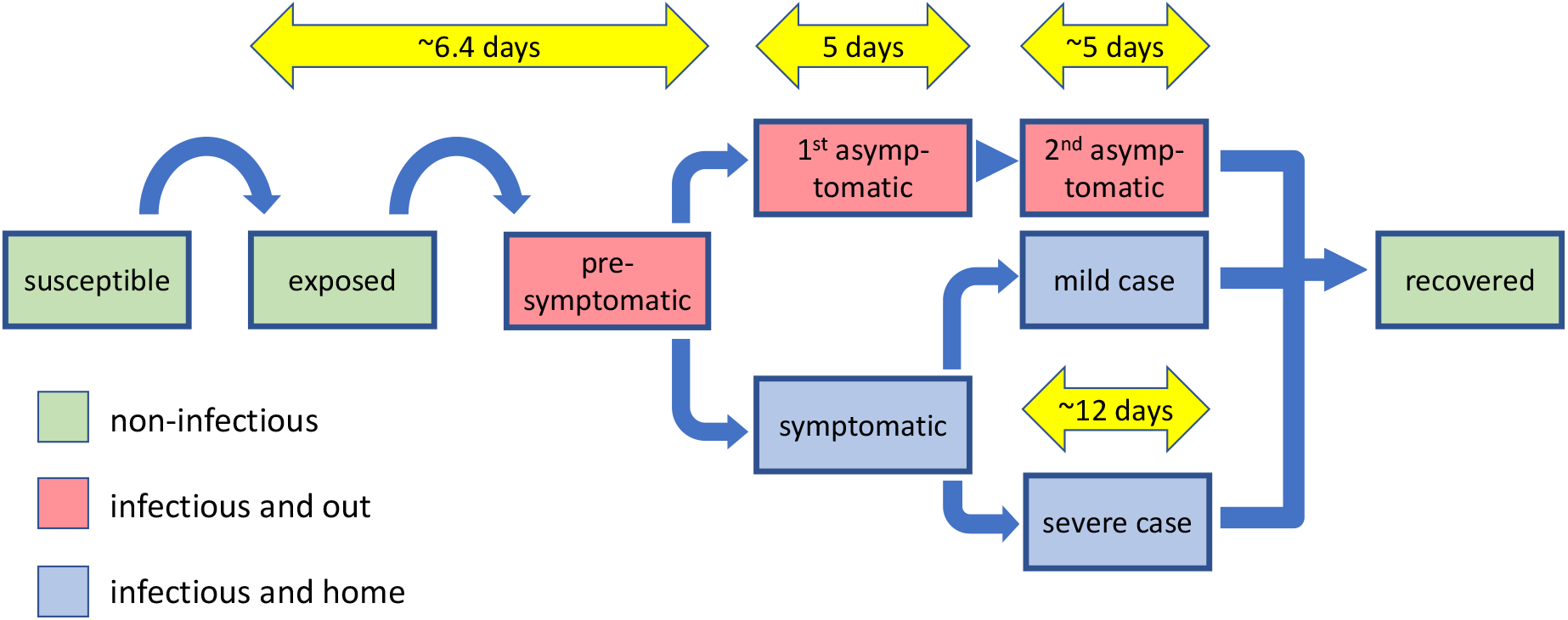
Progression of COVID-19 infection in individuals

### Infection Dynamics

Infection can be transmitted from infectious to susceptible individuals as they go about their daily activities. Let *p_idw_* denote the probability that susceptible individual *i* becomes infected on day *d* by transmission route *w*, where *w* ∈ {*h, t,f,m*} indicates transmission within the household, at toilets, in the food line, or as individuals move about the camp, respectively. The probability that susceptible individual *i* becomes infected on day *d* is thus

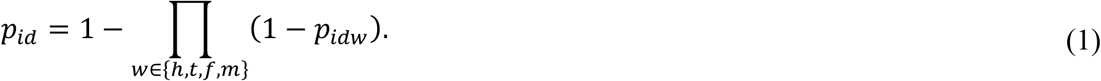

We lack detailed information on how people use space in Moria or any other refugee camp. Therefore, we did not model movement explicitly, but instead calculated the *p_idw_*s for each individual given its expected activities on each day. This reduces the computational time for simulations.

#### Infection within the household

On each day, each infectious individual infects each susceptible individual in the same household with probability *p_h_*. Thus, if individual *i* shares a household with *h_cid_* infectious individuals on day *d*, then

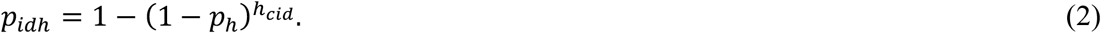

#### Infection at toilets

We assumed that every individual visits the toilet nearest its household 3 times each day, and must always wait in line. If a susceptible individual is in front of or behind an infectious individual in the toilet line, the susceptible individual becomes infected with probability *p_t_*. Thus, the probability that susceptible individual *i* becomes infected in the toilet line on day *d* is

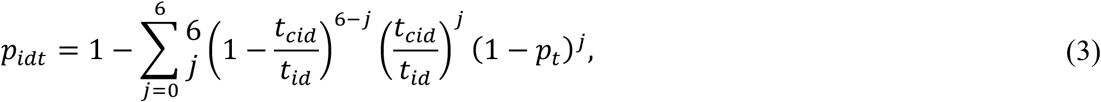

where *t_cid_* and *t_id_* are the numbers of infectious individuals and of all individuals, respectively, that share a toilet with individual *i* on day *d*.

#### Infection in the food line

The food line forms 3 times each day. We assumed that only individuals without symptoms (*i.e*., susceptible, exposed, pre-symptomatic, asymptomatic, or recovered) attend food lines. Food is delivered to individuals with symptoms by others, without interaction (*e.g*., food might be left outside homes). Each individual without symptoms attends the food line once per day on 3 out of 4 days. On other occasions, food is brought to that individual by another individual without additional interactions. For example, food might be brought by a member of the same household, or by a neighbour with whom the individual would otherwise interact (see below). If an individual attends the food line, it interacts with two individuals behind it and two individuals in front of it in the line. Because food lines in Moria are extremely dense,^13,14^ this may be conservative. If a susceptible individual interacts with an infectious individual in the food line, the susceptible individual becomes infected with probability *p_f_*. Thus, the probability that susceptible individual *i* becomes infected in the food line on day *d* is

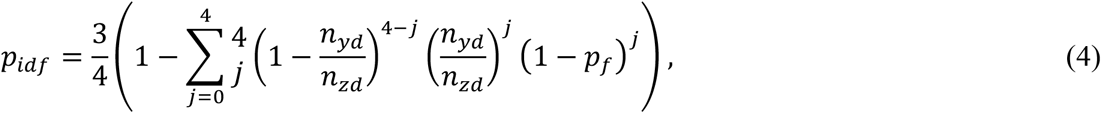

where *n_yd_* is the number of infectious individuals without symptoms (*i.e*., pre-symptomatic and asymptomatic) in the camp on day *d*, and *n_zd_* is the total number of individuals without symptoms in the camp on day *d*.

#### Infection as individuals move about the camp

Individuals move about outside their households, and interact with individuals from other households as they move. We assumed that each individual occupies a circular home range centred on its household, and uses all parts of its home range equally. Two individuals may interact if their home ranges overlap. If individuals *i* and *j* have home ranges with radii *r_i_* and *r_j_*, respectively, and the distance between their households is *d_ij_*, then the area of overlap in their home ranges is

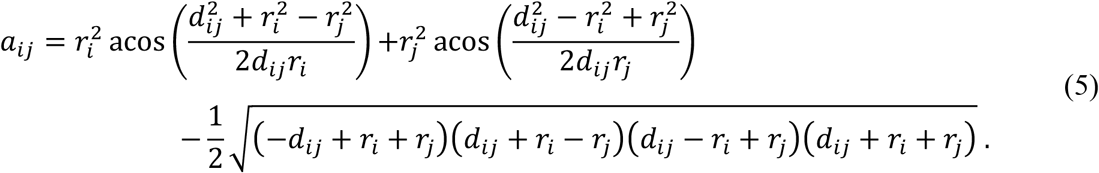

The proportion of time that individuals *i* and *j* spend together in the area of overlap is

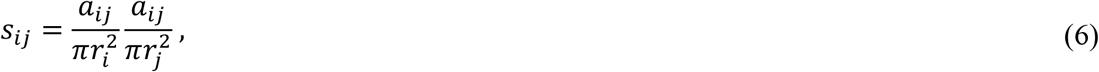

and the relative encounter rate between individuals *i* and *j* is

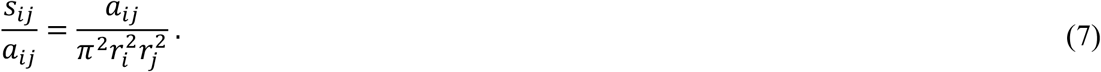

Equation (7) means that individuals encounter each other more frequently if they co-occupy a small area than if they co-occupy a large area for the same amount of time. To obtain the interaction rate between individuals *i* and *j* from the relative encounter rate, we scaled by a factor *g_ij_* to account for ethnicity or country of origin. In particular, *g_ij_ =* 1 if individuals *i* and *j* have the same background, and *g_ij_ =* 0.2 otherwise. Furthermore, we scaled the interaction rate such that two individuals with the same background that share an identical home range with a radius of *r_s_* interact on average once each day. The parameter *r_s_* allows us to scale the mean interaction rate in the population independent of the area that people occupy outside their homes. After scaling, the daily rate of interaction between individuals *i* and *j* is

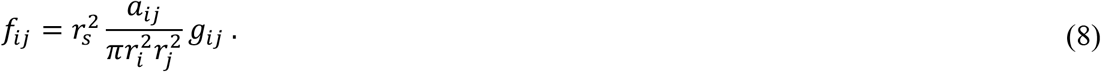

We assumed that only individuals without symptoms interact in their home ranges. Thus, the rate at which individual *i* interacts with infected individuals in its home range on day *d* is

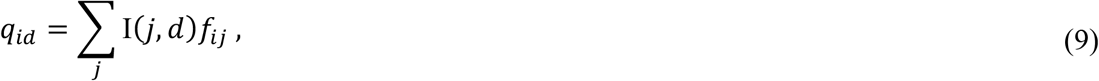

where *I*(*j,d) =* 1 if individual *j* is pre-symptomatic or asymptomatic on day *d* and *I*(*j,d) =* 0 otherwise. The summation in equation (9) runs over all individuals in the model that do not share a household with individual *i*. The probability that susceptible individual *i* becomes infected on day *d* while moving about its home range is thus

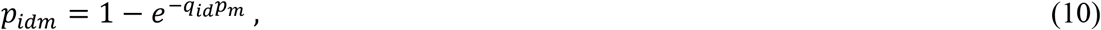

where *p_m_* is the probability of transmission when a susceptible individual interacts with an infectious individual.

#### Assigning parameter values

The probabilities that COVID-19 is transmitted among individuals in different settings are not well-understood. Therefore, in the body of this paper we report results for high-transmission (*p_h_* = 0.18*,p_t_ =* 0.051 *,p_f_=* 0.23, *p_m_ =* 0.0085) and low-transmission (*p_h_ =* 0.0397, *p_t_ =* 0.0067*,p_f_=* 0.0397, *p_m_* = 0.0060) scenarios. These values are derived from the literature^19,21,22,25^ in the supplementary information. We also know very little about how people use space or interact in Moria or in other refugee camps. Therefore, we modelled high- and low-movement and high- and low interaction scenarios. In the high-movement scenario, we assumed that males over 10 years old use home ranges with radius 0.2 (*i.e*., 200 m), and that males under 10 years old and all females use home ranges with radius 0.05. In the low movement scenario, we assumed that males over 10 years old use home ranges with radius 0.1, and all others use home ranges with radius 0.02. In the high-interaction scenario, we set *r_s_* so that the average individual in the camp interacts with 20 others per day (*i.e., r, = 0.0226* and *r, =* 0.0202 in high- the low-movement scenarios, respectively). In the low-interaction scenario, we set *r_s_* so that the average individual in the camp interacts with 5 others per day (*i.e., r_s_ =* 0.0113 and *r_s_ =* 0.0101 in high- the low-movement scenarios, respectively).

For each combination of transmission, movement, and interaction scenario, we estimated the basic reproduction number R_0_ by conducting 10^4^ simulations. In each simulation, we allowed a randomly selected individual in the population to become infected, and we counted the number of individuals infected by this index case (table S12). In low-transmission scenarios, R_0_ ranged from 4.02 to 4.64 depending on the movement and interaction rates. This is slightly higher than in Chinese cities before interventions^32^. In high-transmission scenarios, R_0_ ranged from 14.44 to 15.38, in line with estimates from the Diamond Princess before interventions^33^. With shared food lines, shared toilets, and a population density of >20,000 km^−2^, Moria may be more similar to a cruise ship than to a city, but with more crowded housing and less sanitation. We believe our low- and high-transmission scenarios represent plausible upper and lower bounds for the transmission potential of COVID-19 in Moria.

### Interventions

We modelled four different interventions that might be imposed on the baseline model, alone and in combinations: sectoring, face mask use, remove-and-isolate, and lockdown.

#### Sectoring

The camp in our baseline model has a single food line where transmission can occur among individuals from any parts of the camp. This facilitates the rapid spread of infection. A plausible intervention would be to divide the camp into sectors with separate food lines, and to require individuals to use the food line closest to their homes. To simulate such an intervention, we divided the camp into *n* sectors, each with its own food line. These sectors form a 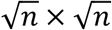 grid over the camp. We replaced equation (4) with

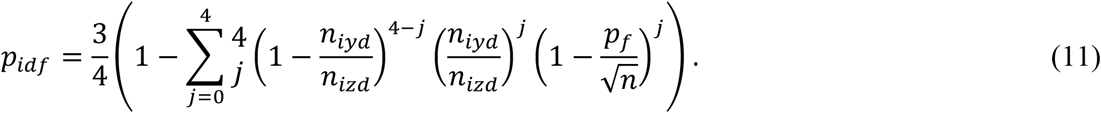

Here *n_iyd_* is the number of infectious individuals without symptoms (*i.e*., pre-symptomatic and asymptomatic) served by the same food line as individual *i* on day *d*, and *n_izd_* is the total number of individuals without symptoms served by the same food line as individual *i* on day *d*. Rescaling the transmission probability by 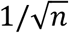 accounts for the fact that shorter lines have shorter waiting times. We conducted simulations with *n* ∈ {4, 16, 144} to study how the number of sectors affects COVID-19 epidemics.

#### Face mask use

Behavioral changes such as using personal protective equipment, frequent handwashing, and maintaining safe distances from others may reduce the risk of COVID-19 transmission. In Moria, there is approximately one tap per 42 people, so frequent handwashing (*e.g*., greater than 10× per day, as in^23^) may be impossible. Due to the high population density, maintaining safe distances from others may also be difficult or impossible.^5^ However, people in Moria have been provided with face masks. To simulate the use of face masks, we scaled the odds of transmission per interaction in food lines, in toilet lines, and during movement about the camp by a factor of 0.32 following Jefferson and colleagues.^23^

#### Remove-and-isolate

Managers of some populations, including Moria, have planned interventions in which people with COVID-19 infections and their households will be removed from populations and kept in isolation until the infected people have recovered. By isolating entire households, managers aim to remove asymptomatic and pre-symptomatic individuals from the population, and to ensure that carers are not separated from their families. To simulate a remove-and-isolate intervention, we conducted simulations in which in each individual with symptoms (*i.e*., symptomatic, mild case, or severe case) is detected with probability *b* on each day. If an individual with symptoms is detected, that individual and its household are removed from the camp. Individuals removed from the camp can infect or become infected by others in their households following equation (2), but cannot infect or become infected by individuals in other households by any transmission route. We assumed that individuals are returned to the camp 7 days after they have recovered, or if they do not become infected, 7 days after the last infected person in their household has recovered. We simulated remove-and-isolate interventions with *b* ∈ {1, 0.5, 0.25}. These capture interventions in which symptomatic individuals and their households are removed on average on the 1^st^, 2^nd^, or 4^th^ day of symptoms.

#### Lockdown

Some countries have attempted to limit the spread of COVID-19 by requiring people to stay in or close to their homes.^11^ This intervention has sometimes been called “lockdown.” We simulated lockdowns in which most individuals are restricted to home ranges with radius *r_l_* around their households, except when visiting shared toilets or food lines. We assumed that a proportion *ν_l_* of the population violates the lockdown. Thus, for each individual in the population, we set their home range to *r_l_* with probability (1- *ν_l_*). Otherwise, we set their home range to 0.2 in the high-movement scenario or to 0.1 in the low movement scenario. We simulated interventions with (*r_l_, ν_l_*) ∈ {(0.005, 0.05), (0.01, 0.1), (0.02, 0.2)} to study lockdowns that are more or less restrictive and strictly enforced.

### Simulations

In each simulation, we initialised the model population and camp structure as described above, and we randomly selected one individual to enter the exposed state. We simulated the epidemic by iterating days, and we tracked the disease state of each individual over time. We ran each simulation until all individuals in the population were either susceptible or recovered, at which point the epidemic had ended. If fewer than 20 individuals became infected, we recorded that an epidemic had been averted. If the epidemic was not averted, then we recorded the maximum number of infected individuals, the time to peak infection, and the proportion of the population that became infected in each simulation. For remove-and-isolate interventions, we also recorded the peak number of individuals in isolation to assess feasibility.

## Data Availability

Original codes for models presented in this study are currently available from the corresponding author. Upon publication of the study, codes and data will be made freely available through the Open Science Foundation (https://osf.io/).

## Acknowledgments

The authors thank Folashade B Agusto for comments and the University of Manchester IT Services for access to high throughput computing on the HTCondor system. RTG was partially supported by Engineering and Physical Sciences Research Council grant number EP/M506436.

## Supplementary Materials

### Estimating transmission probabilities for the high- and low-transmission scenarios

#### High-transmission scenarios

For the initial high-transmission scenario (HT1), we estimated the daily transmission probability within households, *p_h_*, using data from Danis and colleagues.^25^ Danis and colleagues reported that 8 of 10 people who shared an apartment in a French chalet for four days with one infectious individual subsequently became infected. Thus, we estimated *p_h_* = 0.33 by solving 1 − (1 −*p_h_*)^4^ = 8/10.

We estimated transmission rates per interaction using data from Liu and colleagues.^21^ Liu and colleagues reported a total of 43 secondary infections among 126 attendees at 8 meals, each with one infectious individual present. We assumed that meals lasted 2 h and that the transmission rate was constant over time. Thus, the probability of transmission in an interaction lasting *m* minutes is

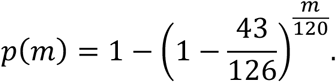

We assumed that interactions in food lines, toilet lines, and while moving about the camp lasted for 150 min, 30 min, and 5 min respectively. Therefore, *p_f_=p*(150) = 0.407, *p_t_ =p*(30) *=* 0.099, and *p_f_=p*(5) = 0.017.

The transmission parameters in HT1 result in R_0_s in that range from 22.1 to 23.8 depending on the movement and interaction rates in the model population. These are higher than have been observed for COVID-19 in any real population. Therefore, we created a second high-transmission scenario (HT2) in which we reduced the rate of transmission in each transmission route by a factor of 0.5. Thus,

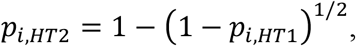

where *p_i,HT1_* and *p_i,HT2_* are the transmission probabilities for transmission route *i* in high-transmission scenarios HT1 and HT2, respectively. In HT2*, p_h_ =* 0.18*,p_f_=* 0.23*,p_t_ =* 0.051, and *p_f_ =* 0.0085. Without interventions R_0_ ranges from 14.44 to 15.38, in line with estimates from the Diamond Princess before interventions.^33^ In the body of this paper, we present results from HT2. Results from HT1 are presented in supplementary tables S1-S12.

#### Low-transmission scenarios

For the initial low-transmission scenario (LT1), we estimated the daily transmission probability within households using data from Li and colleagues.^22^ Li and colleagues studied the households of 105 COVID-19 patients who were hospitalised in China between 1 January and 20 February 2020. Household members were exposed to infection until patients were hospitalised, and Li and colleagues recorded the proportion of household members that became infected.

Members of households occupying isoboxes or tents in the Moria refugee camp may be in closer contact for longer periods than members of Chinese households. Therefore, we assumed that the transmission rates among household members in Moria would be similar to the transmission rates between spouses in Chinese households, who may be in closer contact than other household members.

Li and colleagues reported that 25 of 90 spouses of infectious individuals became infected. However, spouses in Li and colleagues’ data were exposed to their infectious partners for multiple days, and our model is parameterised on daily transmission probabilities. Therefore, we estimated the days of exposure for spouses in Li and colleagues’ data set, and used this and the total infection rate to estimate the daily transmission probability. Li and colleagues reported that 12 patients were hospitalised on days 0 or 1 of symptoms, 34 were hospitalised on days 2-5 of symptoms, and 59 were hospitalised on days 7-11 of symptoms. Fourteen patients self-isolated in their homes from the onset of symptoms and there was no transmission from these patients to their households. We do not know on which days the patients that self-isolated were hospitalised, so we assumed that they were divided proportionally between the group that was hospitalised on days 2-5 and the group that was hospitalised on days 7-11. We assumed that every patient became infectious three days before the appearance of symptoms^24^ and remained infections until hospitalisation. We do not know the exact day on which patients were hospitalised, so we assumed that all patients were hospitalised on the middle day for their groups. We solved

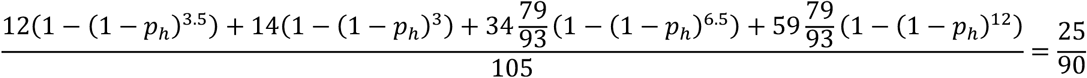

for *p_h_* to obtain an estimated daily transmission probability within households of 0.0397. Because the Moria population has smaller homes, less sanitary conditions (*e.g*., no washing facilities in homes), and poorer background health than the population Li and colleagues studied, this estimate may be conservative.

We set the transmission probability between individuals that interact in food lines, *p_f_*, equal to *p_h_*. This is reasonable because food lines in Moria are dense and people wait in food lines for up to 3 h per visit.^13,14^ We set the transmission probability between individuals that interact in toilet lines to 1 − (1 −*p_f_*)^1/6^ = 0.0067 to reflect an estimated 30 min waiting time in toilet lines. We set the transmission rate per interaction during movement in the camp to *p_m_* = 0.006 following Shen and colleagues.^19^ Shen and colleagues reported that 3 of 473 of attendees at three parties with 2 infectious individuals became infected. It is unlikely that the 2 infectious individuals interacted with all of the other attendees at each party. Thus, Shen and colleagues’ estimate may be conservative as a per-interaction transmission probability.

The transmission parameters in LT1 result in R_0_s in that range from 4.02 to 4.64, slightly higher than those observed in Chinese cities before interventions.^32^ This is plausible because conditions in Moria are likely to favour transmission more than those in Chinese cities.

Because they are estimated from different sources, the relative rates of transmission among transmission routes (*e.g*., transmission in toilet lines relative to transmission in casual interactions during daily activities) differ between LT1 and the high-transmission scenarios. To show that differences between our high- and low-transmission scenarios are due to overall transmission and not to differences in the relative transmission rates among transmission routes, we created a second low-transmission scenario (LT2) by rescaling the transmission rates in HT1. In particular, we set

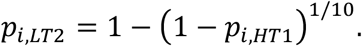

Thus, in LT2, *p_h_* = 0.039, *p_f_=* 0.051, *p_t_=* 0.010, and *p_f_=* 0.0017, and without interventions R_0_ ranges from 4.32 to 4.51 depending on the movement and interaction rates in the model population.

In the body of this paper, we report results for LT1. Figure S1 shows that the qualitative results presented in figures 1 and 2 also hold under LT2.

**Figure S1.**
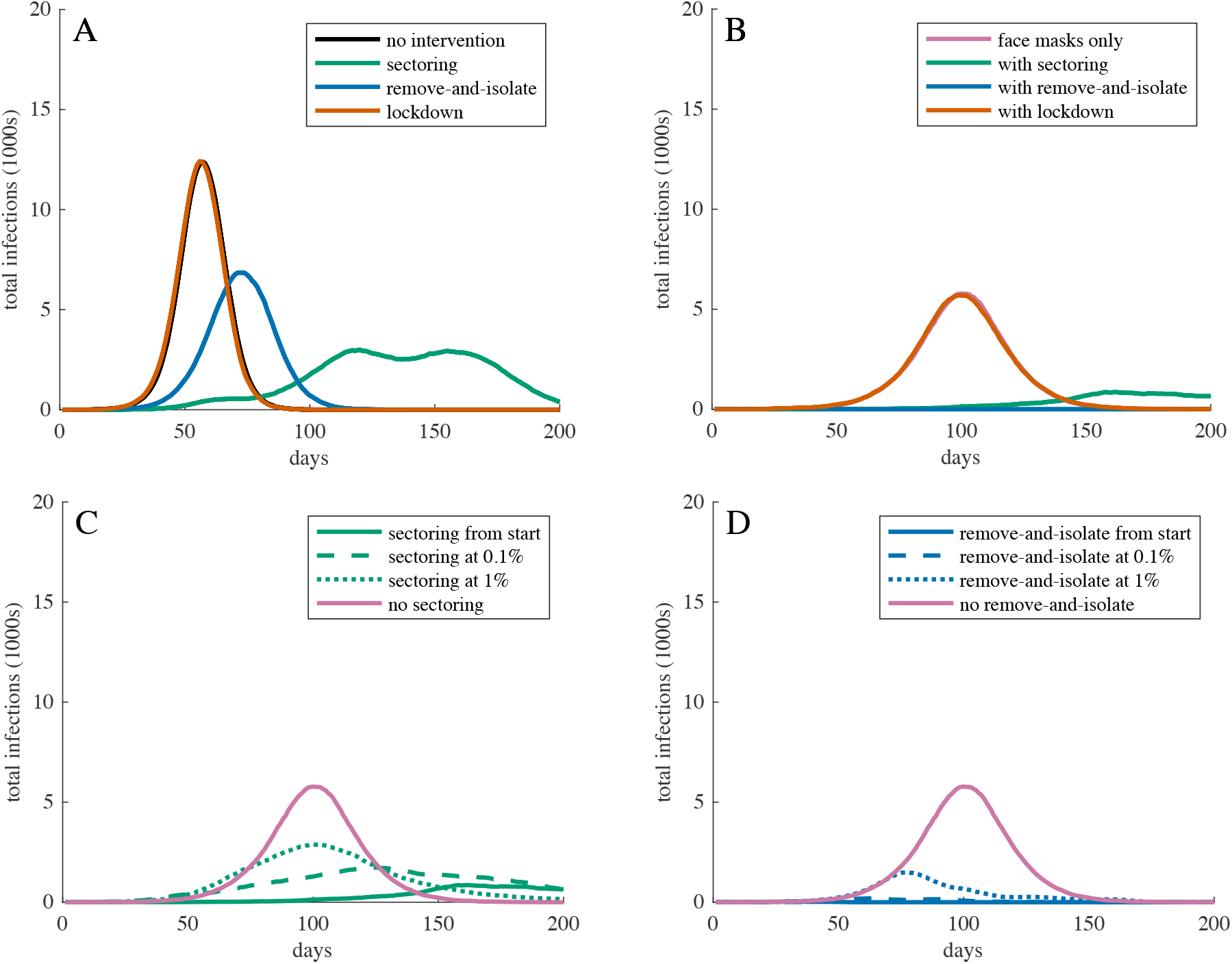
Total infections over time for COVID-19 epidemics with different interventions in populations with low movement, high interaction rates, and low transmission (LT2) probabilities. Intervention intensities are as in figures 1 and 2. Curves show the most representative simulation (*i.e*., the simulation with the peak infection and peak infection date closest to the median) for the corresponding intervention. (A) shows the effect of each intervention alone (compare to figure 1A). (B) shows the effect of each intervention when face masks are also in use (compare to figure 1C). (C) shows the effect of sectoring beginning after infection is detected in the camp (compare to figure 2A). (D) shows the effect of remove-and-isolate beginning after infection has been detected in the camp (compare to figure 2C).

## Supplementary Tables

Supplementary tables S1-S11 report summary statistics for COVID-19 introductions into the model population in scenarios without (S1) or with (S2-S11) interventions. Each row represents 200 simulations. The peak and total proportions of individuals infected, time to peak infection, and peak population in isolation are reported as medians with interquartile ranges. Epidemics adverted is the proportion of simulations in which fewer than 20 individuals became infected. Transmission probabilities for the low- and high-transmission scenarios are presented in the supplementary information, and assumptions of the low- and high-movement and low- and high-interaction scenarios are explained in the Methods. Rows highlighted in pink represent the scenarios reported in the body of the paper.

**Table S1.**

| Interventions | Transmission | Movement | Interaction | Peak proportion infected | Time to peak infection | Total proportion infected | Epidemics averted | Peak population in isolation |
| --- | --- | --- | --- | --- | --- | --- | --- | --- |
| No intervention | Low (LT1) | Low | Low | 0.57 (0.57-0.58) | 64 (60-70) | 0.97 (0.97-0.97) | 0.05 |  |
|  |  |  | High | 0.67 (0.66-0.67) | 55 (52-59) | 0.98 (0.98-0.98) | 0.03 |  |
|  |  | High | Low | 0.58 (0.57-0.58) | 65 (61-71) | 0.97 (0.97-0.97) | 0.05 |  |
|  |  |  | High | 0.68 (0.67-0.68) | 55 (51-59) | 0.98 (0.98-0.98) | 0.02 |  |
|  | High (HT2) | Low | Low | 0.98 (0.98-0.98) | 27 (25-27) | >0.99 | <0.01 |  |
|  |  |  | High | 0.98 (0.98-0.98) | 25 (25-27) | >0.99 | <0.01 |  |
|  |  | High | Low | 0.98 (0.98-0.98) | 26 (25-28) | >0.99 | <0.01 |  |
|  |  |  | High | 0.98 (0.98-0.98) | 26 (25-27) | >0.99 | <0.01 |  |
|  | High (HT1) | Low | Low | >0.99 | 21 (20-22) | >0.99 | <0.01 |  |
|  |  |  | High | >0.99 | 20 (19-21) | >0.99 | <0.01 |  |
|  |  | High | Low | >0.99 | 21 (20-22) | >0.99 | <0.01 |  |
|  |  |  | High | >0.99 | 20 (19-21) | >0.99 | <0.01 |  |

**Table S2.**

| Interventions | Transmission | Movement | Interaction | Peak proportion infected | Time to peak infection | Total proportion infected | Epidemics averted | Peak population in isolation |
| --- | --- | --- | --- | --- | --- | --- | --- | --- |
| Face masks | Low (LT1) | Low | Low | 0.23 (0.22-0.23) | 119 (111-129) | 0.80 (0.80-0.81) | 0.22 |  |
|  |  |  | High | 0.31 (0.31-0.32) | 96 (89-106) | 0.87 (0.87-0.88) | 0.17 |  |
|  |  | High | Low | 0.23 (0.23-0.24) | 116 (108-126) | 0.80 (0.80-0.81) | 0.21 |  |
|  |  |  | High | 0.32 (0.32-0.33) | 92 (84-101) | 0.87 (0.87-0.88) | 0.20 |  |
|  | High (HT2) | Low | Low | 0.88 (0.88-0.89) | 37 (35-39) | >0.99 | <0.01 |  |
|  |  |  | High | 0.90 (0.89-0.90) | 36 (34-38) | >0.99 | <0.01 |  |
|  |  | High | Low | 0.89 (0.88-0.89) | 37 (36-39) | >0.99 | <0.01 |  |
|  |  |  | High | 0.90 (0.90-0.90) | 35 (34-37) | >0.99 | <0.01 |  |
|  | High (HT1) | Low | Low | 0.97 (0.97-0.97) | 27 (26-29) | >0.99 | <0.01 |  |
|  |  |  | High | 0.97 (0.97-0.97) | 27 (26-28) | >0.99 | <0.01 |  |
|  |  | High | Low | 0.97 (0.97-0.97) | 27 (26-29) | >0.99 | <0.01 |  |
|  |  |  | High | 0.97 (0.97-0.98) | 26 (25-28) | >0.99 | <0.01 |  |

**Table S3.**

| Interventions | Transmission | Movement | Interaction | Peak proportion infected | Time to peak infection | Total proportion infected | Epidemics averted | Peak population in isolation |
| --- | --- | --- | --- | --- | --- | --- | --- | --- |
| 4 sectors | Low (LT1) | Low | Low | 0.25 (0.24-0.27) | 103 (95-111) | 0.94 (0.94-0.94) | 0.08 |  |
|  |  |  | High | 0.34 (0.32-0.37) | 78 (73-84) | 0.97 (0.97-0.97) | 0.08 |  |
|  |  | High | Low | 0.30 (0.28-0.32) | 91 (83-101) | 0.94 (0.94-0.94) | 0.07 |  |
|  |  |  | High | 0.43 (0.40-0.46) | 68 (62-73) | 0.97 (0.97-0.97) | 0.05 |  |
|  | High (HT2) | Low | Low | 0.60 (0.54-0.66) | 43 (40-45) | >0.99 | <0.01 |  |
|  |  |  | High | 0.67 (0.65-0.71) | 38 (35-40) | >0.99 | <0.01 |  |
|  |  | High | Low | 0.68 (0.64-0.73) | 40 (37-42) | >0.99 | <0.01 |  |
|  |  |  | High | 0.79 (0.76-0.83) | 34 (33-36) | >0.99 | <0.01 |  |
|  | High (HT1) | Low | Low | 0.73 (0.68-0.77) | 33 (32-36) | >0.99 | <0.01 |  |
|  |  |  | High | 0.83 (0.79-0.86) | 30 (28-31) | >0.99 | <0.01 |  |
|  |  | High | Low | 0.83 (0.78-0.86) | 31 (29-32) | >0.99 | <0.01 |  |
|  |  |  | High | 0.91 (0.89-0.93) | 27 (26-28) | >0.99 | <0.01 |  |
| 16 sectors | Low (LT1) | Low | Low | 0.13 (0.13-0.15) | 138 (117-158) | 0.91 (0.91-0.91) | 0.07 |  |
|  |  |  | High | 0.20 (0.19-0.22) | 98 (86-111) | 0.96 (0.96-0.96) | 0.05 |  |
|  |  | High | Low | 0.18 (0.17-0.20) | 108 (95-127) | 0.91 (0.91-0.91) | 0.10 |  |
|  |  |  | High | 0.31 (0.28-0.34) | 73 (68-84) | 0.96 (0.96-0.96) | 0.05 |  |
|  | High (HT2) | Low | Low | 0.34 (0.32-0.39) | 61 (54-70) | >0.99 | <0.01 |  |
|  |  |  | High | 0.41 (0.39-0.47) | 51 (45-57) | >0.99 | <0.01 |  |
|  |  | High | Low | 0.43 (0.40-0.49) | 50 (46-59) | >0.99 | <0.01 |  |
|  |  |  | High | 0.55 (0.52-0.62) | 43 (38-47) | >0.99 | <0.01 |  |
|  | High (HT1) | Low | Low | 0.45 (0.43-0.52) | 46 (41-53) | >0.99 | <0.01 |  |
|  |  |  | High | 0.54 (0.51-0.62) | 40 (35-44) | >0.99 | <0.01 |  |
|  |  | High | Low | 0.55 (0.53-0.62) | 39 (36-44) | >0.99 | <0.01 |  |
|  |  |  | High | 0.71 (0.67-0.77) | 34 (31-36) | >0.99 | <0.01 |  |
| 144 sectors | Low (LT1) | Low | Low | 0.08 (0.07-0.09) | 210 (173-252) | 0.88 (0.88-0.88) | 0.14 |  |
|  |  |  | High | 0.15 (0.14-0.17) | 113 (98-136) | 0.95 (0.95-0.96) | 0.06 |  |
|  |  | High | Low | 0.14 (0.13-0.15) | 130 (111-162) | 0.88 (0.88-0.89) | 0.10 |  |
|  |  |  | High | 0.27 (0.26-0.30) | 78 (71-91) | 0.95 (0.95-0.95) | 0.06 |  |
|  | High (HT2) | Low | Low | 0.19 (0.17-0.21) | 101 (82-119) | >0.99 | <0.01 |  |
|  |  |  | High | 0.27 (0.26-0.30) | 68 (58-79) | >0.99 | <0.01 |  |
|  |  | High | Low | 0.33 (0.30-0.36) | 64 (57-75) | >0.99 | <0.01 |  |
|  |  |  | High | 0.48 (0.45-0.53) | 47 (42-51) | >0.99 | <0.01 |  |
|  | High (HT1) | Low | Low | 0.26 (0.24-0.29) | 73 (60-82) | >0.99 | <0.01 |  |
|  |  |  | High | 0.36 (0.34-0.41) | 53 (44-60) | >0.99 | <0.01 |  |
|  |  | High | Low | 0.46 (0.42-0.50) | 47 (43-53) | >0.99 | <0.01 |  |
|  |  |  | High | 0.62 (0.59-0.67) | 37 (34-41) | >0.99 | <0.01 |  |

**Table S4.**

| Interventions | Transmission | Movement | Interaction | Peak proportion infected | Time to peak infection | Total proportion infected | Epidemics averted | Peak population in isolation |
| --- | --- | --- | --- | --- | --- | --- | --- | --- |
| Remove and isolate on ~day 4 | Low (LT1) | Low | Low | 0.31 (0.30-0.32) | 84 (79-91) | 0.83 (0.83-0.84) | 0.28 | 0.43 (0.42-0.44) |
|  |  |  | High | 0.46 (0.45-0.47) | 65 (61-70) | 0.90 (0.90-0.90) | 0.20 | 0.59 (0.58-0.60) |
|  |  | High | Low | 0.31 (0.30-0.33) | 82 (77-90) | 0.83 (0.83-0.84) | 0.26 | 0.43 (0.42-0.45) |
|  |  |  | High | 0.47 (0.46-0.48) | 62 (58-67) | 0.90 (0.90-0.90) | 0.17 | 0.60 (0.59-0.61) |
|  | High (HT2) | Low | Low | 0.97 (0.96-0.97) | 28 (27-29) | >0.99 | <0.01 | 0.92 (0.92-0.92) |
|  |  |  | High | 0.97 (0.97-0.97) | 27 (26-28) | >0.99 | 0.02 | 0.93 (0.92-0.93) |
|  |  | High | Low | 0.97 (0.97-0.97) | 27 (26-29) | >0.99 | <0.01 | 0.92 (0.92-0.92) |
|  |  |  | High | 0.97 (0.97-0.97) | 27 (26-28) | >0.99 | <0.01 | 0.93 (0.93-0.93) |
|  | High (HT1) | Low | Low | >0.99 | 21 (20-22) | >0.99 | <0.01 | 0.95 (0.95-0.95) |
|  |  |  | High | >0.99 | 21 (20-22) | >0.99 | <0.01 | 0.95 (0.95-0.95) |
|  |  | High | Low | >0.99 | 22 (21-22) | >0.99 | <0.01 | 0.95 (0.94-0.95) |
|  |  |  | High | >0.99 | 21 (20-22) | >0.99 | <0.01 | 0.95 (0.95-0.95) |
| Remove and isolate on ~day 2 | Low (LT1) | Low | Low | 0.23 (0.22-0.24) | 94 (85-104) | 0.77 (0.76-0.78) | 0.43 | 0.36 (0.34-0.38) |
|  |  |  | High | 0.39 (0.38-0.40) | 70 (65-76) | 0.87 (0.86-0.87) | 0.27 | 0.57 (0.55-0.58) |
|  |  | High | Low | 0.23 (0.22-0.24) | 92 (83-103) | 0.77 (0.76-0.78) | 0.43 | 0.36 (0.34-0.38) |
|  |  |  | High | 0.41 (0.40-0.42) | 66 (62-72) | 0.87 (0.87-0.87) | 0.24 | 0.58 (0.57-0.59) |
|  | High (HT2) | Low | Low | 0.96 (0.96-0.96) | 29 (27-30) | >0.99 | 0.01 | 0.95 (0.94-0.95) |
|  |  |  | High | 0.97 (0.96-0.97) | 27 (26-29) | >0.99 | 0.02 | 0.95 (0.95-0.95) |
|  |  | High | Low | 0.96 (0.96-0.96) | 28 (27-29) | >0.99 | <0.01 | 0.95 (0.94-0.95) |
|  |  |  | High | 0.97 (0.97-0.97) | 27 (26-28) | >0.99 | 0.02 | 0.95 (0.95-0.95) |
|  | High (HT1) | Low | Low | >0.99 | 22 (21-23) | >0.99 | <0.01 | 0.97 (0.97-0.97) |
|  |  |  | High | >0.99 | 21 (20-22) | >0.99 | <0.01 | 0.97 (0.97-0.97) |
|  |  | High | Low | >0.99 | 22 (21-23) | >0.99 | <0.01 | 0.97 (0.97-0.97) |
|  |  |  | High | >0.99 | 21 (20-22) | >0.99 | <0.01 | 0.97 (0.97-0.97) |
| Remove and isolate on ~day 1 | Low (LT1) | Low | Low | 0.18 (0.16-0.20) | 104 (93-111) | 0.71 (0.70-0.73) | 0.40 | 0.30 (0.27-0.32) |
|  |  |  | High | 0.35 (0.34-0.36) | 72 (67-81) | 0.85 (0.84-0.85) | 0.40 | 0.54 (0.52-0.54) |
|  |  | High | Low | 0.19 (0.17-0.20) | 102 (91-110) | 0.72 (0.70-0.73) | 0.47 | 0.31 (0.28-0.33) |
|  |  |  | High | 0.36 (0.35-0.37) | 68 (63-76) | 0.85 (0.84-0.85) | 0.32 | 0.56 (0.54-0.57) |
|  | High (HT2) | Low | Low | 0.95 (0.95-0.95) | 29 (28-31) | >0.99 | 0.01 | 0.95 (0.95-0.95) |
|  |  |  | High | 0.96 (0.96-0.96) | 28 (27-29) | >0.99 | 0.01 | 0.96 (0.96-0.96) |
|  |  | High | Low | 0.95 (0.95-0.95) | 29 (28-30) | >0.99 | <0.01 | 0.95 (0.95-0.95) |
|  |  |  | High | 0.96 (0.96-0.96) | 28 (27-29) | >0.99 | <0.01 | 0.96 (0.96-0.96) |
|  | High (HT1) | Low | Low | >0.99 | 22 (21-23) | >0.99 | <0.01 | 0.97 (0.97-0.97) |
|  |  |  | High | >0.99 | 21 (21-22) | >0.99 | <0.01 | 0.98 (0.97-0.98) |
|  |  | High | Low | >0.99 | 22 (21-23) | >0.99 | <0.01 | 0.97 (0.97-0.98) |
|  |  |  | High | >0.99 | 21 (20-22) | >0.99 | <0.01 | 0.98 (0.97-0.97) |

**Table S5.**

| Interventions | Transmission | Movement | Interaction | Peak proportion infected | Time to peak infection | Total proportion infected | Epidemics averted | Peak population in isolation |
| --- | --- | --- | --- | --- | --- | --- | --- | --- |
| Loose lockdown | Low (LT1) | Low | Low | 0.58 (0.57-0.58) | 65 (61-69) | 0.97 (0.97-0.97) | 0.02 |  |
|  |  |  | High | 0.67 (0.67-0.68) | 55 (53-60) | 0.99 (0.98-0.99) | 0.02 |  |
|  |  | High | Low | 0.58 (0.58-0.59) | 64 (61-67) | 0.97 (0.97-0.97) | 0.06 |  |
|  |  |  | High | 0.69 (0.68-0.69) | 54 (52-58) | 0.99 (0.99-0.99) | 0.03 |  |
|  | High (HT2) | Low | Low | 0.98 (0.98-0.98) | 26 (25-28) | >0.99 | <0.01 |  |
|  |  |  | High | 0.98 (0.98-0.98) | 26 (25-27) | >0.99 | <0.01 |  |
|  |  | High | Low | 0.98 (0.98-0.98) | 27 (25-28) | >0.99 | <0.01 |  |
|  |  |  | High | 0.98 (0.98-0.98) | 25 (24-27) | >0.99 | <0.01 |  |
|  | High (HT1) | Low | Low | >0.99 | 21 (20-22) | >0.99 | <0.01 |  |
|  |  |  | High | >0.99 | 20 (19-21) | >0.99 | <0.01 |  |
|  |  | High | Low | >0.99 | 21 (20-22) | >0.99 | <0.01 |  |
|  |  |  | High | >0.99 | 20 (19-21) | >0.99 | <0.01 |  |
| Moderate lockdown | Low (LT1) | Low | Low | 0.57 (0.57-0.58) | 65 (61-68) | 0.97 (0.97-0.97) | 0.04 |  |
|  |  |  | High | 0.66 (0.65-0.66) | 57 (54-61) | 0.98 (0.98-0.99) | 0.04 |  |
|  |  | High | Low | 0.58 (0.58-0.59) | 64 (61-68) | 0.97 (0.97-0.97) | 0.06 |  |
|  |  |  | High | 0.68 (0.67-0.68) | 55 (53-59) | 0.99 (0.99-0.99) | 0.02 |  |
|  | High (HT2) | Low | Low | 0.98 (0.98-0.98) | 26 (25-28) | >0.99 | <0.01 |  |
|  |  |  | High | 0.98 (0.98-0.98) | 26 (25-27) | >0.99 | <0.01 |  |
|  |  | High | Low | 0.98 (0.98-0.98) | 26 (25-28) | >0.99 | <0.01 |  |
|  |  |  | High | 0.98 (0.98-0.98) | 26 (24-27) | >0.99 | <0.01 |  |
|  | High (HT1) | Low | Low | >0.99 | 21 (20-22) | >0.99 | <0.01 |  |
|  |  |  | High | >0.99 | 20 (20-21) | >0.99 | <0.01 |  |
|  |  | High | Low | >0.99 | 21 (20-22) | >0.99 | <0.01 |  |
|  |  |  | High | >0.99 | 20 (19-21) | >0.99 | <0.01 |  |
| Tight lockdown | Low (LT1) | Low | Low | 0.57 (0.56-0.57) | 66 (63-70) | 0.97 (0.97-0.97) | 0.06 |  |
|  |  |  | High | 0.63 (0.62-0.63) | 59 (56-63) | 0.98 (0.98-0.98) | 0.04 |  |
|  |  | High | Low | 0.58 (0.57-0.58) | 66 (63-71) | 0.97 (0.97-0.97) | 0.04 |  |
|  |  |  | High | 0.64 (0.63-0.64) | 58 (55-63) | 0.98 (0.98-0.98) | 0.02 |  |
|  | High (HT2) | Low | Low | 0.98 (0.98-0.98) | 26 (25-28) | >0.99 | <0.01 |  |
|  |  |  | High | 0.98 (0.98-0.98) | 26 (25-27) | >0.99 | <0.01 |  |
|  |  | High | Low | 0.98 (0.98-0.98) | 26 (25-28) | >0.99 | <0.01 |  |
|  |  |  | High | 0.98 (0.98-0.98) | 26 (25-27) | >0.99 | <0.01 |  |
|  | High (HT1) | Low | Low | >0.99 | 21 (20-22) | >0.99 | <0.01 |  |
|  |  |  | High | >0.99 | 21 (20-22) | >0.99 | <0.01 |  |
|  |  | High | Low | >0.99 | 21 (20-22) | >0.99 | <0.01 |  |
|  |  |  | High | >0.99 | 21 (20-21) | >0.99 | <0.01 |  |

**Table S6.**

| Interventions | Transmission | Movement | Interaction | Peak proportion infected | Time to peak infection | Total proportion infected | Epidemics averted | Peak population in isolation |
| --- | --- | --- | --- | --- | --- | --- | --- | --- |
| 4 sectors with face masks | Low (LT1) | Low | Low | 0.073 (0.067-0.084) | 217 (190-244) | 0.68 (0.68-0.69) | 0.37 |  |
|  |  |  | High | 0.14 (0.13-0.15) | 145 (130-165) | 0.82 (0.82-0.82) | 0.23 |  |
|  |  | High | Low | 0.098 (0.088-0.11) | 180 (159-198) | 0.68 (0.68-0.69) | 0.31 |  |
|  |  |  | High | 0.19 (0.18-0.21) | 119 (106-133) | 0.82 (0.82-0.83) | 0.22 |  |
|  | High (HT2) | Low | Low | 0.41 (0.39-0.44) | 62 (59-65) | >0.99 | <0.01 |  |
|  |  |  | High | 0.48 (0.45-0.52) | 55 (51-58) | >0.99 | <0.01 |  |
|  |  | High | Low | 0.49 (0.46-0.55) | 59 (56-62) | >0.99 | <0.01 |  |
|  |  |  | High | 0.58 (0.55-0.62) | 50 (47-53) | >0.99 | <0.01 |  |
|  | High (HT1) | Low | Low | 0.53 (0.50-0.61) | 45 (42-47) | >0.99 | <0.01 |  |
|  |  |  | High | 0.65 (0.61-0.70) | 39 (36-42) | >0.99 | <0.01 |  |
|  |  | High | Low | 0.66 (0.61-0.70) | 41 (39-44) | >0.99 | <0.01 |  |
|  |  |  | High | 0.75 (0.72-0.78) | 36 (34-38) | >0.99 | <0.01 |  |
| 16 sectors with face masks | Low (LT1) | Low | Low | 0.037 (0.030-0.043) | 277 (195-341) | 0.50 (0.47-0.52) | 0.42 |  |
|  |  |  | High | 0.090 (0.082-0.10) | 167 (137-207) | 0.77 (0.76-0.78) | 0.26 |  |
|  |  | High | Low | 0.054 (0.049-0.060) | 199 (166-247) | 0.54 (0.53-0.56) | 0.42 |  |
|  |  |  | High | 0.13 (0.13-0.14) | 129 (108-149) | 0.78 (0.77-0.78) | 0.25 |  |
|  | High (HT2) | Low | Low | 0.22 (0.21-0.25) | 92 (81-110) | >0.99 | <0.01 |  |
|  |  |  | High | 0.28 (0.26-0.30) | 76 (66-84) | >0.99 | 0.01 |  |
|  |  | High | Low | 0.27 (0.25-0.31) | 80 (71-92) | >0.99 | <0.01 |  |
|  |  |  | High | 0.36 (0.34-0.39) | 62 (55-70) | >0.99 | <0.01 |  |
|  | High (HT1) | Low | Low | 0.32 (0.29-0.36) | 65 (58-76) | >0.99 | <0.01 |  |
|  |  |  | High | 0.39 (0.36-0.42) | 53 (48-62) | >0.99 | <0.01 |  |
|  |  | High | Low | 0.39 (0.37-0.43) | 53 (48-64) | >0.99 | <0.01 |  |
|  |  |  | High | 0.51 (0.48-0.57) | 44 (40-50) | >0.99 | <0.01 |  |
| 144 sectors with face masks | Low (LT1) | Low | Low | 0.009 (0.002-0.014) | 177 (91-294) | 0.15 (0.01-0.23) | 0.60 |  |
|  |  |  | High | 0.063 (0.056-0.068) | 218 (176-260) | 0.71 (0.70-0.72) | 0.28 |  |
|  |  | High | Low | 0.032 (0.025-0.035) | 228 (164-288) | 0.38 (0.34-0.40) | 0.60 |  |
|  |  |  | High | 0.11 (0.11-0.12) | 135 (114-166) | 0.73 (0.72-0.74) | 0.29 |  |
|  | High (HT2) | Low | Low | 0.11 (0.10-0.12) | 174 (143-208) | 0.99 (0.98-0.99) | <0.01 |  |
|  |  |  | High | 0.17 (0.16-0.19) | 106 (87-126) | >0.99 | <0.01 |  |
|  |  | High | Low | 0.19 (0.17-0.21) | 103 (89-121) | >0.99 | 0.01 |  |
|  |  |  | High | 0.31 (0.38-0.34) | 71 (61-80) | >0.99 | <0.01 |  |
|  | High (HT1) | Low | Low | 0.17 (0.15-0.19) | 108 (92-130) | >0.99 | <0.01 |  |
|  |  |  | High | 0.25 (0.23-0.27) | 76 (62-88) | >0.99 | <0.01 |  |
|  |  | High | Low | 0.29 (0.26-0.32) | 70 (62-84) | >0.99 | <0.01 |  |
|  |  |  | High | 0.43 (0.40-0.47) | 51 (45-58) | >0.99 | <0.01 |  |

**Table S7.**
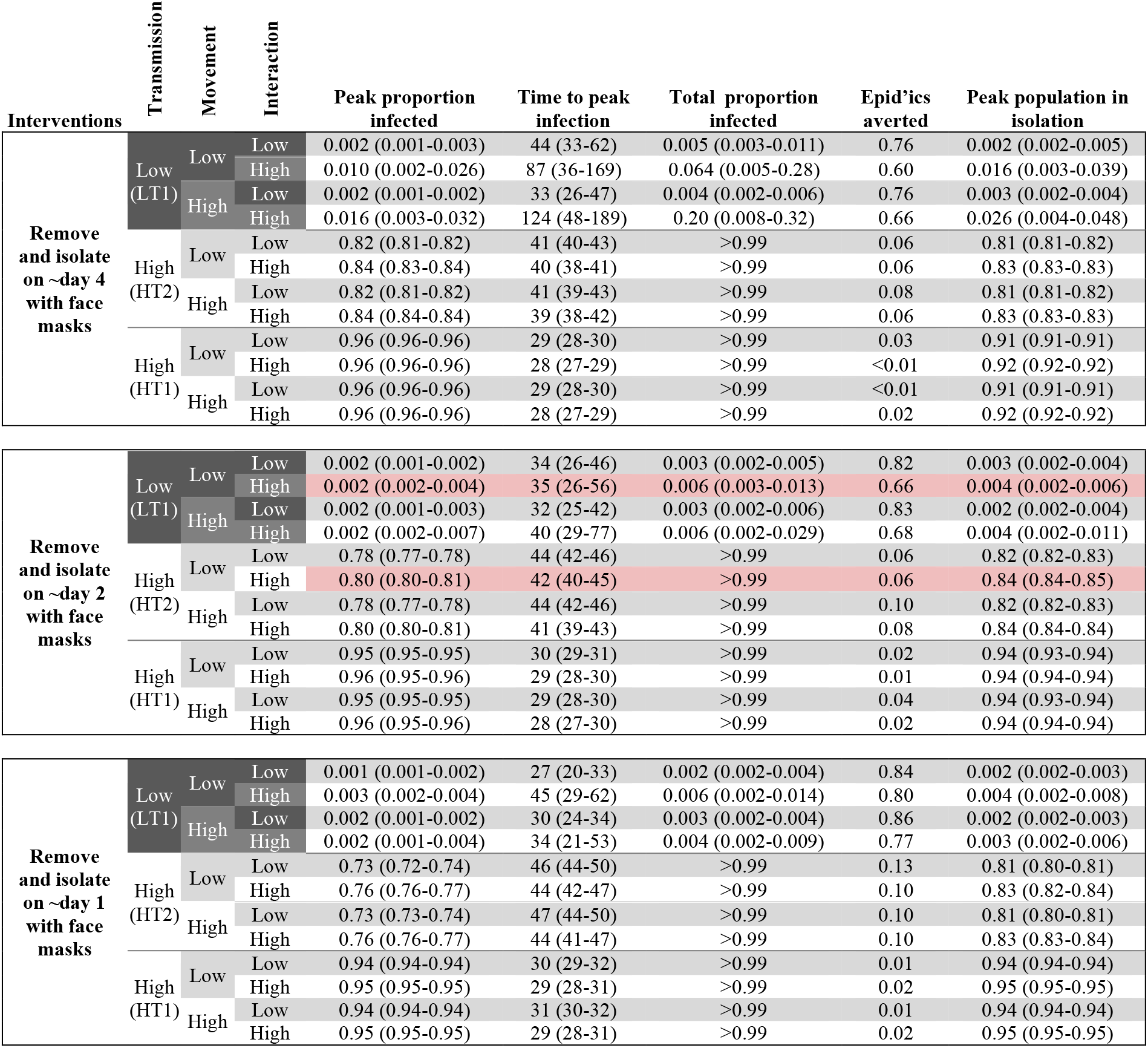

**Table S8.**

| Interventions | Transmission | Movement | Interaction | Peak proportion infected | Time to peak infection | Total proportion infected | Epidemics averted | Peak population in isolation |
| --- | --- | --- | --- | --- | --- | --- | --- | --- |
| Loose lockdown with face masks | Low (LT1) | Low | Low | 0.23 (0.22-0.23) | 120 (110-132) | 0.80 (0.80-0.81) | 0.24 |  |
|  |  |  | High | 0.31 (0.31-0.32) | 97 (90-104) | 0.88 (0.87-0.88) | 0.14 |  |
|  |  | High | Low | 0.23 (0.23-0.24) | 119 (108-129) | 0.81 (0.81-0.82) | 0.22 |  |
|  |  |  | High | 0.33 (0.32-0.33) | 95 (87-104) | 0.89 (0.88-0.89) | 0.14 |  |
|  | High (HT2) | Low | Low | 0.89 (0.88-0.89) | 37 (35-39) | >0.99 | <0.01 |  |
|  |  |  | High | 0.90 (0.89-0.90) | 36 (34-37) | >0.99 | <0.01 |  |
|  |  | High | Low | 0.89 (0.88-0.89) | 37 (35-39) | >0.99 | <0.01 |  |
|  |  |  | High | 0.90 (0.90-0.90) | 36 (34-37) | >0.99 | <0.01 |  |
|  | High (HT1) | Low | Low | 0.97 (0.97-0.97) | 27 (26-28) | >0.99 | <0.01 |  |
|  |  |  | High | 0.97 (0.97-0.97) | 27 (25-28) | >0.99 | <0.01 |  |
|  |  | High | Low | 0.97 (0.97-0.97) | 27 (26-29) | >0.99 | <0.01 |  |
|  |  |  | High | 0.97 (0.97-0.98) | 26 (25-28) | >0.99 | <0.01 |  |
| Moderate lockdown with face masks | Low (LT1) | Low | Low | 0.22 (0.22-0.23) | 122 (112-135) | 0.80 (0.80-0.81) | 0.19 |  |
|  |  |  | High | 0.30 (0.29-0.30) | 102 (95-112) | 0.87 (0.87-0.88) | 0.14 |  |
|  |  | High | Low | 0.23 (0.22-0.24) | 118 (108-125) | 0.81 (0.81-0.82) | 0.22 |  |
|  |  |  | High | 0.31 (0.31-0.32) | 99 (92-108) | 0.88 (0.88-0.89) | 0.14 |  |
|  | High (HT2) | Low | Low | 0.89 (0.88-0.89) | 37 (36-39) | >0.99 | <0.01 |  |
|  |  |  | High | 0.90 (0.89-0.90) | 36 (34-37) | >0.99 | <0.01 |  |
|  |  | High | Low | 0.89 (0.88-0.89) | 37 (35-39) | >0.99 | <0.01 |  |
|  |  |  | High | 0.90 (0.90-0.90) | 36 (34-38) | >0.99 | <0.01 |  |
|  | High (HT1) | Low | Low | 0.97 (0.97-0.97) | 27 (26-28) | >0.99 | <0.01 |  |
|  |  |  | High | 0.97 (0.97-0.97) | 26 (26-28) | >0.99 | <0.01 |  |
|  |  | High | Low | 0.97 (0.97-0.97) | 28 (26-29) | >0.99 | <0.01 |  |
|  |  |  | High | 0.97 (0.97-0.98) | 27 (25-28) | >0.99 | <0.01 |  |
| Tight lockdown with face masks | Low (LT1) | Low | Low | 0.21 (0.21-0.22) | 124 (115-137) | 0.80 (0.79-0.80) | 0.24 |  |
|  |  |  | High | 0.26 (0.26-0.27) | 112 (102-122) | 0.85 (0.85-0.86) | 0.22 |  |
|  |  | High | Low | 0.22 (0.22-0.23) | 121 (113-134) | 0.81 (0.80-0.81) | 0.16 |  |
|  |  |  | High | 0.27 (0.27-0.28) | 107 (100-119) | 0.86 (0.86-0.86) | 0.27 |  |
|  | High (HT2) | Low | Low | 0.88 (0.88-0.89) | 37 (36-39) | >0.99 | <0.01 |  |
|  |  |  | High | 0.89 (0.89-0.90) | 36 (35-38) | >0.99 | <0.01 |  |
|  |  | High | Low | 0.89 (0.88-0.89) | 37 (35-39) | >0.99 | <0.01 |  |
|  |  |  | High | 0.90 (0.89-0.90) | 36 (34-38) | >0.99 | <0.01 |  |
|  | High (HT1) | Low | Low | 0.97 (0.97-0.97) | 27 (26-28) | >0.99 | <0.01 |  |
|  |  |  | High | 0.97 (0.97-0.97) | 26 (26-28) | >0.99 | <0.01 |  |
|  |  | High | Low | 0.97 (0.97-0.97) | 27 (26-29) | >0.99 | <0.01 |  |
|  |  |  | High | 0.97 (0.97-0.97) | 27 (26-28) | >0.99 | <0.01 |  |

**Table S9.**
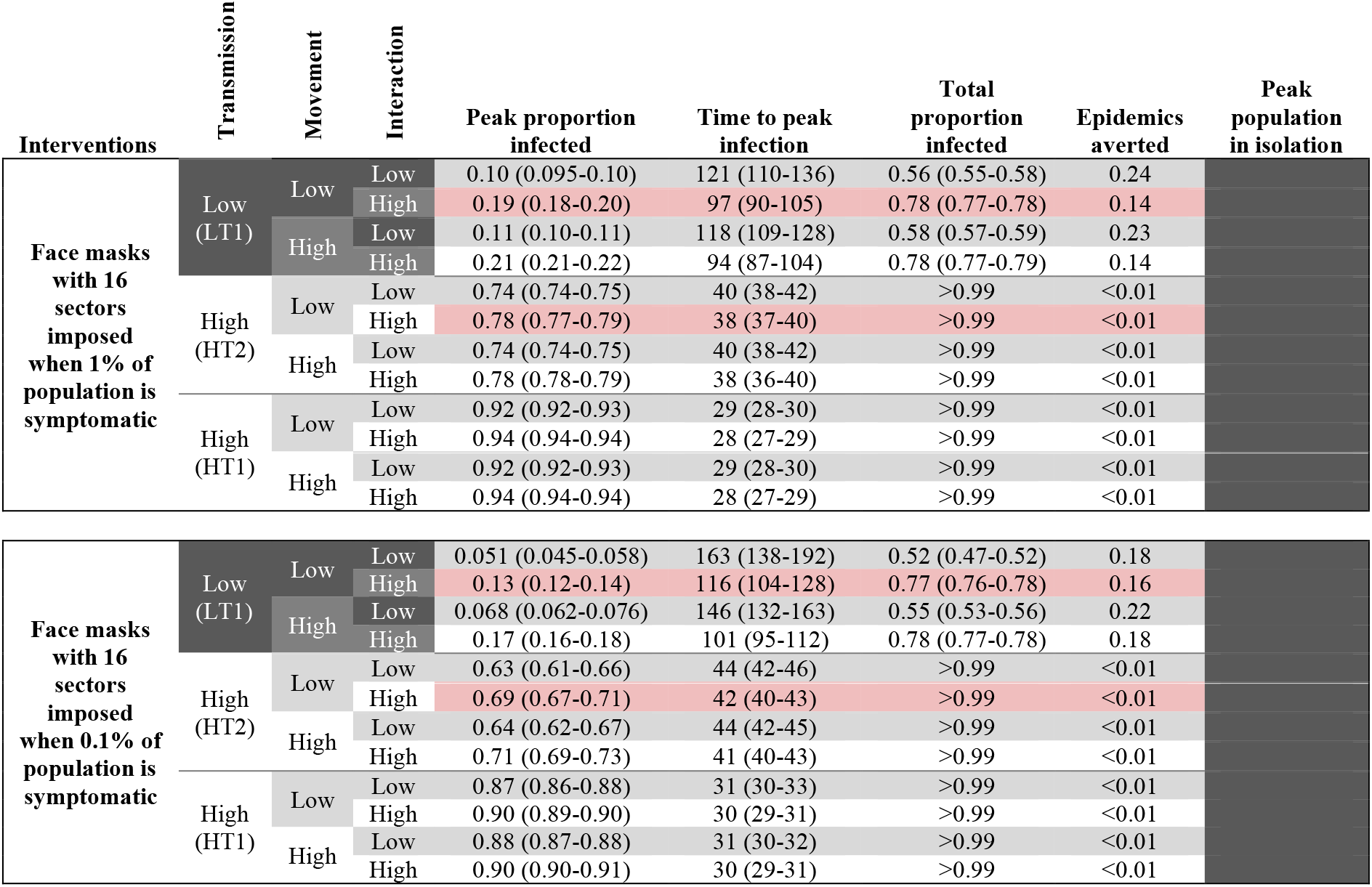

| Interventions | Transmission | Movement | Interaction | Peak proportion infected | Time to peak infection | Total proportion infected | Epidemics averted | Peak population in isolation |
| --- | --- | --- | --- | --- | --- | --- | --- | --- |
| Face masks with 16 sectors imposed when 1% of population is symptomatic | Low (LT1) | Low | Low | 0.10 (0.095-0.10) | 121 (110-136) | 0.56 (0.55-0.58) | 0.24 |  |
|  |  |  | High | 0.19 (0.18-0.20) | 97 (90-105) | 0.78 (0.77-0.78) | 0.14 |  |
|  |  | High | Low | 0.11 (0.10-0.11) | 118 (109-128) | 0.58 (0.57-0.59) | 0.23 |  |
|  |  |  | High | 0.21 (0.21-0.22) | 94 (87-104) | 0.78 (0.77-0.79) | 0.14 |  |
|  | High (HT2) | Low | Low | 0.74 (0.74-0.75) | 40 (38-42) | >0.99 | <0.01 |  |
|  |  |  | High | 0.78 (0.77-0.79) | 38 (37-40) | >0.99 | <0.01 |  |
|  |  | High | Low | 0.74 (0.74-0.75) | 40 (38-42) | >0.99 | <0.01 |  |
|  |  |  | High | 0.78 (0.78-0.79) | 38 (36-40) | >0.99 | <0.01 |  |
|  | High (HT1) | Low | Low | 0.92 (0.92-0.93) | 29 (28-30) | >0.99 | <0.01 |  |
|  |  |  | High | 0.94 (0.94-0.94) | 28 (27-29) | >0.99 | <0.01 |  |
|  |  | High | Low | 0.92 (0.92-0.93) | 29 (28-30) | >0.99 | <0.01 |  |
|  |  |  | High | 0.94 (0.94-0.94) | 28 (27-29) | >0.99 | <0.01 |  |
| Face masks with 16 sectors imposed when 0.1% of population is symptomatic | Low (LT1) | Low | Low | 0.051 (0.045-0.058) | 163 (138-192) | 0.52 (0.47-0.52) | 0.18 |  |
|  |  |  | High | 0.13 (0.12-0.14) | 116 (104-128) | 0.77 (0.76-0.78) | 0.16 |  |
|  |  | High | Low | 0.068 (0.062-0.076) | 146 (132-163) | 0.55 (0.53-0.56) | 0.22 |  |
|  |  |  | High | 0.17 (0.16-0.18) | 101 (95-112) | 0.78 (0.77-0.78) | 0.18 |  |
|  | High (HT2) | Low | Low | 0.63 (0.61-0.66) | 44 (42-46) | >0.99 | <0.01 |  |
|  |  |  | High | 0.69 (0.67-0.71) | 42 (40-43) | >0.99 | <0.01 |  |
|  |  | High | Low | 0.64 (0.62-0.67) | 44 (42-45) | >0.99 | <0.01 |  |
|  |  |  | High | 0.71 (0.69-0.73) | 41 (40-43) | >0.99 | <0.01 |  |
|  | High (HT1) | Low | Low | 0.87 (0.86-0.88) | 31 (30-33) | >0.99 | <0.01 |  |
|  |  |  | High | 0.90 (0.89-0.90) | 30 (29-31) | >0.99 | <0.01 |  |
|  |  | High | Low | 0.88 (0.87-0.88) | 31 (30-32) | >0.99 | <0.01 |  |
|  |  |  | High | 0.90 (0.90-0.91) | 30 (29-31) | >0.99 | <0.01 |  |

**Table S10.**

| Interventions | Transmission | Movement | Interaction | Peak proportion infected | Time to peak infection | Total proportion infected | Epidemics averted | Peak population in isolation |
| --- | --- | --- | --- | --- | --- | --- | --- | --- |
| Face masks with remove-and-isolate on ~day 2 starting when 1% of population is symptomatic | Low (LT1) | Low | Low | 0.059 (0.056-0.064) | 89 (82-104) | 0.17 (0.16-0.19) | 0.24 | 0.091 (0.086-0.097) |
|  |  |  | High | 0.086 (0.082-0.093) | 76 (69-88) | 0.30 (0.28-0.31) | 0.10 | 0.13 (0.13-0.14) |
|  |  | High | Low | 0.061 (0.057-0.066) | 87 (77-97) | 0.17 (0.16-0.19) | 0.26 | 0.093 (0.087-0.10) |
|  |  |  | High | 0.097 (0.091-0.10) | 74 (68-83) | 0.34 (0.31-0.35) | 0.18 | 0.15 (0.14-0.16) |
|  | High (HT2) | Low | Low | 0.81 (0.81-0.82) | 38 (37-40) | >0.99 | <0.01 | 0.85 (0.85-0.86) |
|  |  |  | High | 0.84 (0.83-0.84) | 37 (36-39) | >0.99 | <0.01 | 0.87 (0.87-0.87) |
|  |  | High | Low | 0.81 (0.81-0.82) | 38 (36-40) | >0.99 | <0.01 | 0.85 (0.85-0.86) |
|  |  |  | High | 0.84 (0.83-0.84) | 37 (35-39) | >0.99 | <0.01 | 0.87 (0.87-0.87) |
|  | High (HT1) | Low | Low | 0.96 (0.96-0.96) | 28 (26-29) | >0.99 | <0.01 | 0.94 (0.94-0.95) |
|  |  |  | High | 0.96 (0.96-0.97) | 27 (26-28) | >0.99 | <0.01 | 0.95 (0.95-0.95) |
|  |  | High | Low | 0.96 (0.96-0.96) | 28 (27-29) | >0.99 | <0.01 | 0.94 (0.94-0.95) |
|  |  |  | High | 0.97 (0.96-0.97) | 27 (26-28) | >0.99 | <0.01 | 0.95 (0.95-0.95) |
| Face masks with remove-and-isolate on ~day 2 starting when 0.1% of population is symptomatic | Low (LT1) | Low | Low | 0.007 (0.006-0.009) | 59 (49-81) | 0.020 (0.015-0.026) | 0.24 | 0.011 (0.009-0.014) |
|  |  |  | High | 0.013 (0.010-0.017) | 66 (52-83) | 0.058 (0.038-0.11) | 0.18 | 0.021 (0.016-0.027) |
|  |  | High | Low | 0.007 (0.006-0.009) | 58 (50-69) | 0.020 (0.015-0.027) | 0.21 | 0.011 (0.009-0.014) |
|  |  |  | High | 0.016 (0.013-0.021) | 72 (58-102) | 0.097 (0.061-0.14) | 0.14 | 0.026 (0.021-0.033) |
|  | High (HT2) | Low | Low | 0.78 (0.78-0.79) | 40 (39-42) | >0.99 | <0.01 | 0.83 (0.82-0.83) |
|  |  |  | High | 0.80 (0.80-0.81) | 39 (37-40) | >0.99 | <0.01 | 0.84 (0.84-0.85) |
|  |  | High | Low | 0.78 (0.78-0.79) | 40 (39-42) | >0.99 | <0.01 | 0.83 (0.82-0.83) |
|  |  |  | High | 0.81 (0.80-0.81) | 38 (37-40) | >0.99 | <0.01 | 0.85 (0.84-0.85) |
|  | High (HT1) | Low | Low | 0.95 (0.95-0.95) | 28 (27-30) | >0.99 | <0.01 | 0.94 (0.94-0.94) |
|  |  |  | High | 0.96 (0.96-0.96) | 28 (26-29) | >0.99 | <0.01 | 0.94 (0.94-0.95) |
|  |  | High | Low | 0.95 (0.95-0.95) | 28 (27-29) | >0.99 | <0.01 | 0.94 (0.94-0.94) |
|  |  |  | High | 0.96 (0.96-0.96) | 28 (27-29) | >0.99 | <0.01 | 0.94 (0.94-0.95) |

**Table S11.**

| Interventions | Transmission | Movement | Interaction | Peak proportion infected | Time to peak infection | Total proportion infected | Epid'ics averted | Peak population in isolation |
| --- | --- | --- | --- | --- | --- | --- | --- | --- |
| Face masks, sectoring, remove-and-isolate | Low (LT1) | Low | Low | 0.001 (0.001-0.001) | 24 (22-27) | 0.002 (0.002-0.002) | 0.93 | 0.002 (0.002-0.002) |
|  |  |  | High | 0.002 (0.001-0.002) | 32 (19-49) | 0.003 (0.002-0.04) | 0.87 | 0.002 (0.002-0.003) |
|  |  | High | Low | 0.001 (0.001-0.002) | 22 (20-23) | 0.001 (0.001-0.003) | 0.94 | 0.002 (0.001-0.002) |
|  |  |  | High | 0.002 (0.001-0.003) | 30 (24-40) | 0.003 (0.002-0.008) | 0.86 | 0.003 (0.002-0.004) |
|  | High (HT2) | Low | Low | 0.090 (0.075-0.11) | 132 (99-192) | 0.80 (0.62-0.90) | 0.23 | 0.11 (0.091-0.13) |
|  |  |  | High | 0.18 (0.16-0.21) | 98 (83-115) | 0.94 (0.94-0.95) | 0.22 | 0.22 (0.20-0.25) |
|  |  | High | Low | 0.14 (0.13-0.16) | 121 (107-146) | 0.90 (0.89-0.91) | 0.18 | 0.17 (0.15-0.19) |
|  |  |  | High | 0.25 (0.23-0.27) | 78 (70-90) | 0.94 (0.94-0.95) | 0.21 | 0.30 (0.27-0.32) |
|  | High (HT1) | Low | Low | 0.25 (0.23-0.29) | 79 (69-96) | >0.99 | 0.08 | 0.29 (0.26-0.33) |
|  |  |  | High | 0.33 (0.31-0.37) | 62 (53-70) | >0.99 | 0.04 | 0.38 (0.36-0.42) |
|  |  | High | Low | 0.32 (0.30-0.36) | 68 (57-78) | >0.99 | 0.07 | 0.37 (0.33-0.40) |
|  |  |  | High | 0.44 (0.41-0.50) | 48 (45-55) | >0.99 | <0.01 | 0.50 (0.47-0.56) |
| Face masks, sectoring, lockdown | Low (LT1) | Low | Low | 0.020 (0.008-0.026) | 234 (156-311) | 0.32 (0.091-0.40) | 0.52 |  |
|  |  |  | High | 0.060 (0.053-0.065) | 242 (194-294) | 0.75 (0.74-0.76) | 0.26 |  |
|  |  | High | Low | 0.031 (0.026-0.036) | 269 (184-351) | 0.46 (0.42-0.50) | 0.46 |  |
|  |  |  | High | 0.082 (0.076-0.090) | 182 (150-212) | 0.78 (0.77-0.78) | 0.26 |  |
|  | High (HT2) | Low | Low | 0.19 (0.16-0.21) | 113 (94-133) | >0.99 | <0.01 |  |
|  |  |  | High | 0.24 (0.22-0.27) | 91 (76-104) | >0.99 | <0.01 |  |
|  |  | High | Low | 0.22 (0.20-0.25) | 99 (85-117) | >0.99 | <0.01 |  |
|  |  |  | High | 0.29 (0.27-0.32) | 76 (67-85) | >0.99 | <0.01 |  |
|  | High (HT1) | Low | Low | 0.27 (0.24-0.30) | 77 (66-90) | >0.99 | <0.01 |  |
|  |  |  | High | 0.33 (0.31-0.37) | 62 (53-70) | >0.99 | <0.01 |  |
|  |  | High | Low | 0.33 (0.30-0.37) | 65 (58-77) | >0.99 | <0.01 |  |
|  |  |  | High | 0.40 (0.37-0.45) | 55 (48-62) | >0.99 | <0.01 |  |
| Face masks, remove-and-isolate, lockdown | Low (LT1) | Low | Low | 0.002 (0.001-0.002) | 32 (26-43) | 0.003 (0.002-0.004) | 0.80 | 0.003 (0.002-0.003) |
|  |  |  | High | 0.002 (0.001-0.003) | 31 (25-50) | 0.004 (0.002-0.007) | 0.73 | 0.003 (0.002-0.005) |
|  |  | High | Low | 0.002 (0.001-0.002) | 32 (24-38) | 0.003 (0.002-0.005) | 0.85 | 0.003 (0.002-0.004) |
|  |  |  | High | 0.002 (0.001-0.004) | 29 (23-52) | 0.004 (0.002-0.011) | 0.70 | 0.003 (0.002-0.006) |
|  | High (HT2) | Low | Low | 0.78 (0.77-0.78) | 44 (42-46) | >0.99 | 0.10 | 0.82 (0.82-0.83) |
|  |  |  | High | 0.80 (0.80-0.81) | 42 (40-44) | >0.99 | 0.10 | 0.84 (0.84-0.84) |
|  |  | High | Low | 0.78 (0.77-0.79) | 44 (42-46) | >0.99 | 0.10 | 0.82 (0.82-0.83) |
|  |  |  | High | 0.81 (0.80-0.81) | 42 (40-45) | >0.99 | 0.08 | 0.84 (0.84-0.85) |
|  | High (HT1) | Low | Low | 0.95 (0.95-0.95) | 30 (28-31) | >0.99 | 0.04 | 0.94 (0.93-0.94) |
|  |  |  | High | 0.96 (0.95-0.96) | 29 (28-30) | >0.99 | 0.02 | 0.94 (0.94-0.94) |
|  |  | High | Low | 0.95 (0.95-0.96) | 29 (38-31) | >0.99 | <0.01 | 0.94 (0.93-0.94) |
|  |  |  | High | 0.96 (0.96-0.96) | 28 (27-30) | >0.99 | 0.02 | 0.94 (0.94-0.94) |
| Face masks, sectoring, remove-and-isolate, lockdown | Low (LT1) | Low | Low | 0.001 (0.001-0.002) | 23 (19-28) | 0.002 (0.002-0.003) | 0.92 | 0.002 (0.002-0.002) |
|  |  |  | High | 0.002 (0.001-0.002) | 28 (20-43) | 0.003 (0.002-0.005) | 0.88 | 0.002 (0.002-0.003) |
|  |  | High | Low | 0.002 (0.001-0.002) | 26 (21-32) | 0.002 (0.002-0.004) | 0.94 | 0.002 (0.002-0.003) |
|  |  |  | High | 0.002 (0.001-0.002) | 30 (25-43) | 0.003 (0.002-0.006) | 0.84 | 0.002 (0.002-0.004) |
|  | High (HT2) | Low | Low | 0.035 (0.027-0.050) | 74 (51-115) | 0.16 (0.060-0.29) | 0.24 | 0.041 (0.032-0.060) |
|  |  |  | High | 0.14 (0.12-0.15) | 129 (106-160) | 0.94 (0.93-0.95) | 0.16 | 0.16 (0.14-0.18) |
|  |  | High | Low | 0.068 (0.046-0.088) | 121 (79-158) | 0.53 (0.28-0.73) | 0.28 | 0.083 (0.053-0.11) |
|  |  |  | High | 0.18 (0.16-0.20) | 106 (89-124) | 0.95 (0.95-0.95) | 0.16 | 0.21 (0.19-0.24) |
|  | High (HT1) | Low | Low | 0.17 (0.14-0.21) | 97 (75-124) | >0.99 | 0.07 | 0.19 (0.15-0.23) |
|  |  |  | High | 0.27 (0.24-0.30) | 77 (65-86) | >0.99 | 0.04 | 0.30 (0.28-0.34) |
|  |  | High | Low | 0.24 (0.21-0.27) | 84 (73-98) | >0.99 | 0.08 | 0.27 (0.24-0.30) |
|  |  |  | High | 0.34 (0.31-0.38) | 63 (55-71) | >0.99 | 0.06 | 0.38 (0.35-0.42) |

In table S11, the camp is divided into 16 sectors (*n =* 16), remove-and-isolate occurs on average on day 2 (*b =* 2), and lockdown is moderate (*r_l_* = 0.01, *ν_l_* = 0.1).

**Table S12.**

| Movement | Interaction | Low transmission 1 | Low transmission 2 | High Transmission 1 | High Transmission 2 |
| --- | --- | --- | --- | --- | --- |
| Low | Low | 4.02 (6.02)<br>1, 2, 4, 6, 8 | 4.32 (6.67)<br>1, 2, 4, 6, 9 | 22.05 (68.70)<br>10, 16, 21, 27, 37 | 14.48 (34.41)<br>6, 10, 14, 18, 25 |
|  | High | 4.64 (7.83)<br>1, 3, 4, 6, 10 | 4.47 (6.89)<br>1, 3, 4, 6, 9 | 23.83 (88.17)<br>11, 17, 22, 30, 41 | 15.26 (38.97)<br>6, 11, 15, 19, 27 |
| High | Low | 4.05 (6.40)<br>1, 2, 4, 6, 9 | 4.34 (6.72)<br>1, 2, 4, 6, 9 | 22.06 (70.63)<br>10, 16, 21, 27, 38 | 14.44 (35.28)<br>6, 10, 14, 18, 25 |
|  | High | 4.63 (8.10)<br>1, 3, 4, 6, 10 | 4.51 (7.22)<br>1, 3, 4, 6, 9 | 23.79 (91.10)<br>11, 17, 22, 30, 42 | 15.38 (42.20)<br>6, 11, 15, 19, 27 |

Table S12 reports the basic reproduction number R_0_ for COVID-19 in the model population for each scenario combination (*i.e*., transmission rate, movement, and interaction rate) in the absence of intervention. In each cell, the first line reports R_0_ and (in parentheses) the variance of the number of individuals infected by the index case. The second line reports the 5^th^, 25^th^, 50^th^, 75^th^, and 95^th^ percentiles for the number of people infected by the index case.

## References

1. United Nations High Commissioner for Refugees. Global Trends: Forced Displacement in 2018. Geneva; 2019. https://www.unhcr.org/5d08d7ee7.pdf (accessed 16 Jun 2020).

2. Chen T, Wu D, Chen H, Yan W, Yang D, Chen G, et al. Clinical characteristics of 113 deceased patients with coronavirus disease 2019: retrospective study. BMJ. 2020; 368: m1091.

3. Hermans MPJ, Kooistra J, Cannegieter SC, Rosendaal FR, Mook-Kanamori DO, Nemeth B. Healthcare and disease burden among refugees in long-stay refugee camps at Lesbos, Greece. Eur J Epidemiol. 2017; 32(9): 851–4.

4. Bellos A, Mulholland K, O’Brien KL, Qazi SA, Gayer M, Checchi F. The burden of acute respiratory infections in crisis-affected populations: a systematic review. Confl Health. 2010; 4: 3.

5. Kluge HHP, Jakab Z, Bartovic J, D’Anna V, Severoni S. Refugee and migrant health in the COVID-19 response. Lancet. 2020; 395(10232): 1237–9.

6. United Nations High Commissioner for Refugees. Coronavirus Emergency Appeal. Geneva; 2020. http://reporting.unhcr.org/sites/default/files/CQVID-19%20appeal%20-%20REVISED%20-%20FINAL.pdf (accessed 16 Jun 2020).

7. Wasdani KP, Prasad A. The impossibility of social distancing among the urban poor: the case of an Indian slum in the times of COVID-19. Local Environ. 2020; 25(5): 414–8.

8. Truelove S, Abrahim O, Altare C, Lauer SA, Grantz KH, Azman AS, et al. The potential impact of COVID-19 in refugee camps in Bangladesh and beyond: a modeling study. PLOSMed. 2020; 16(6): el003144.

9. Wong JEL, Leo YS, Tan CC. COVID-19 in Singapore-current experience: critical global issues that require attention and action. JAMA. 2020; 323(13): 1243–4.

10. Saez M, Tobias A, Varga D, Barcelo MA. Effectiveness of the measures to flatten the epidemic curve of COVID-19. The case of Spain. Sci Total Environ. 2020; 727: 138761.

11. Anderson RM, Heesterbeek H, Klinkenberg D, Hollingsworth TD. How will country-based mitigation measures influence the course of the COVID-19 epidemic? Lancet. 2020; 395(10228): 931–4.

12. de Berker H. Overcrowding in Moria refugee camp has reached breaking point. Financial Times. 25 Feb 2020. https://www.ft.com/content/013d95d6-54d3-11ea-a1ef-da1721a0541e (accessed 16 Jun 2020).

13. Rankin J, Smith H. EU urged to evacuate asylum seekers from cramped Greek island camps. The Guardian. 24 Mar 2020. https://www.theguardian.com/world/2020/mar/24/eu-urged-to-evacuate-asvlum-seekers-from-cramped-greek-island-camps-coronavirus (accessed 18 June 2020).

14. Smith H. Anger rises in Lesbos over crowded refugee camps. The Guardian. 9 Nov 2017. https://www.theguardian.com/world/2017/nov/09/anger-rises-in-lesbos-over-crowded-refugee-camps (accessed 18 Jun 2020).

15. Fallon K, Grant H. Lesbos coronavirus case sparks fears for refugee camp. The Guardian. 11 Mar 2020. https://www.theguardian.com/global-development/2020/mar/11/lesbos-coronavirus-case-sparks-fears-for-refugee-camp-moria (accessed 16 Jun 2020).

16. Bansal S, Grenfell BT, Meyers LA. When individual behaviour matters: homogeneous and network models in epidemiology. J R Soc Interface. 2007; 4(16): 879–91.

17. Verity R, Okell LC, Dorigatti I, Winskill P, Whittaker C, Imai N, et al. Estimates of the severity of coronavirus disease 2019: a model-based analysis. Lancet Infect Dis. 2020; 60(6): P669–77.

18. Tuite AR, Fisman DN, Greer AL. Mathematical modelling of COVID-19 transmission and mitigation strategies in the population of Ontario, Canada. CMAJ. 2020; 192(19): E497–E505.

19. Shen Y, Xu W, Li C, Handel A, Martinez L, Ling F, et al. A cluster of COVID-19 infections indicating person-to-person transmission among casual contacts from social gatherings: an outbreak case-contact investigation. Open Forum Infect Dis. 2020: ofaa231.

20. Mizumoto K, Kagaya K, Zarebski A, Chowell G. Estimating the asymptomatic proportion of coronavirus disease 2019 (COVID-19) cases on board the Diamond Princess cruise ship, Yokohama, Japan. 2020. Euro Surveill. 2020; 25(10).

21. Liu Y, Eggo RM, Kucharski AJ. Secondary attack rate and superspreading events for SARS-CoV-2. Lancet. 2020; 395(10227): e47.

22. Li W, Zhang B, Lu J, Liu S, Chang Z, Cao P, et al. The characteristics of household transmission of COVID-19. Clin Infect Dis. 2020: ciaa450.

23. Jefferson T, Del Mar C, Dooley L, Ferroni E, Al-Ansary LA, Bawazeer GA, et al. Physical interventions to interrupt or reduce the spread of respiratory viruses: systematic review. BMJ. 2009; 339: b3675.

24. Du Z, Xu X, Wu Y, Wang L, Cowling BJ, Meyers LA. Serial interval of COVID-19 among publicly reported confirmed cases. Emerg Infect Dis. 2020; 26(6): 1341–3.

25. Danis K, Epaulard O, Bénet T, Gaymard A, Campoy S, Bothelo-Nevers E, et al. Cluster of coronavirus disease 2019 (Covid-19) in the French Alps, 2020. Clin Infect Dis. 2020: ciaa424.

26. Chang SL, Harding N, Zacherson C, Cliff OM, Prokopenko M. Modelling transmission and control of the COVID-19 pandemic in Australia. *arXiv* 2020; 2003.10218v3 (preprint).

27. Cai Y, Liu J, Yang H, Chen T, Yu Q, Chen J, et al. COVID-19 discharged patients in Hunan. China: correlation between early features and prognosis. Research Square 2020; published online 20 Mar. DOI:10.21203/rs.3.rs-18121/v1 (preprint).

28. Backer JA, Klinkenberg D, Wallinga J. Incubation period of 2019 novel coronavirus (2019-nCoV) infections among travellers from Wuhan, China, 20-28 January 2020. Euro Surveill. 2020; 25(5).

29. Levita L, Miller JG, Hartman TK, Murphy J, Shevlin M, McBride O, et al. Impact of COVID-19 on the well-being of young people aged 13 to 24 in the UK. 2020. https://drive.google.eom/file/d/1AOc0wCPqv2gfFSQ_DVmw12vrqQK01z0V/view (accessed 16 Jun 2020).

30. Moore RC, Lee A, Hancock JT, Hailey M, Linos E. Experience with social distancing early in the COVID-19 pandemic in the United States: implications for public health messaging. *MedRXiv* 2020; published online 11 Apr. DOI: 10.1101/2020.04.08.20057067 (preprint).

31. Kirkcaldy RD, King BA, Brooks JT. COVID-19 and postinfection immunity: limited evidence, many remaining questions. JAMA. 2020; 323(22): 2245–6.

32. Lin YF, Duan Q, Zhou Y, Yuan T, Li P, Fitzpatrick T, et al. Spread and Impact of COVID-19 in China: A Systematic Review and Synthesis of Predictions From Transmission-Dynamic Models. Front Med (Lausanne). 2020; 7: 321.

33. Rocklov J, Sjodin H, Wilder-Smith A. COVID-19 outbreak on the Diamond Princess cruise ship: estimating the epidemic potential and effectiveness of public health countermeasures. J Travel Med. 2020; 27(3).

